# Concordant and discordant gene expression signatures of twin placentas in the setting of preeclampsia

**DOI:** 10.1101/2025.08.14.25333727

**Authors:** William E. Ackerman, Irina A. Buhimschi, Hongwu Jing, Thomas L. Brown, Guomao Zhao, Catalin S. Buhimschi

**Affiliations:** Department of Obstetrics and Gynecology, University of Illinois College of Medicine Chicago, Chicago, IL 60612, USA; AI.Health4All Center, University of Illinois College of Medicine Chicago, Chicago, IL 60612, USA; Institute for Health Data Science Research, University of Illinois College of Medicine Chicago, Chicago, IL 60612, USA; Department of Neuroscience, Cell Biology and Physiology, Wright State University Boonshoft School of Medicine, Dayton, OH 45435, USA

**Author notes:** **Corresponding Author:** William E. Ackerman IV, MD, Department of Obstetrics and Gynecology, University of Illinois College of Medicine - Chicago, 820 South Wood Street MC 808, Chicago, IL 60612.

**Keywords:** Preeclampsia, Placenta, Twins, Transcriptomics, Next-generation sequencing

## Abstract

**Background:** Multifetal pregnancies are associated with increased risk for preeclampsia (PreE), but the underlying pathogenesis may differ from singleton gestations. It remains unclear whether both placentas in twin pregnancies complicated by PreE exhibit molecular signatures of the disease simultaneously or in isolation.

**Methods:** We performed RNA sequencing on 32 individual placental samples from twin gestations grouped by clinical PreE diagnosis: 24 dichorionic (DT) and 8 monochorionic (MT). Additionally, 10 newly sequenced singleton PreE placentas were analyzed. These were integrated with a benchmark dataset comprising 15 early-onset PreE and 10 control singleton placentas from our prior study (GSE203507), along with 102 samples from two independent studies (GSE114691 and GSE1482410). Differential expression analysis and a machine learning pipeline were used to identify a 98-transcript classification signature for PreE, achieving 97% accuracy in the benchmark dataset.

**Results:** Twin placentas partially aligned with the molecular spectrum of PreE observed in singleton gestations. Signature scoring revealed transcriptional similarities between twin and singleton PreE placentas across varying clinical severities. Intertwin transcriptional divergence was more pronounced in DT than MT samples, regardless of clinical diagnosis. Functional pathway analysis showed consistent dysregulation patterns in PreE, but with notable heterogeneity among twin samples. Some twin placentas without clinical PreE displayed PreE-like transcriptional signatures, suggesting molecular changes may precede clinical manifestation.

**Conclusions:** These results highlight complexity in PreE pathology in twin gestations. In DT pregnancies complicated by PreE, the placentas may be affected unequally, suggesting that PreE in twins may be associated with a single diseased placenta.

## Introduction

Preeclampsia (PreE) is a multisystemic, progressive hypertensive disease of pregnancy and a significant contributor to global maternal and perinatal morbidity and mortality (1). The exact pathoetiology of PreE is not fully understood, but it is broadly accepted that the placenta plays a significant role (2). Specifically, maldevelopment of the uteroplacental circulation can result in inadequate fetal perfusion to support typical development. The resulting tissue stress can trigger the release of placental factors that elicit systemic effects in susceptible maternal systems (3). The pathogenesis of PreE is likely multifactorial, which may explain the observed variability in symptomatology, clinical course (particularly the gestational age at detection), risk factors, and biochemical markers (4, 5).

Multifetal pregnancies are at increased risk for developing PreE (6, 7). In multifetal gestations, PreE tends to occur earlier, is more severe, and results in more adverse outcomes than in singleton gestations (SGs) (8, 9). However, the underlying pathogenesis of PreE in multifetal pregnancies may differ from that in singletons (10). Specifically, twins and high-order multiple pregnancies have greater total placental mass than singletons; this may lead to an excess of circulating anti-angiogenic factors that contribute to the syndrome (11). Studies of placentas from twin pregnancies with PreE revealed fewer pathological lesions related to placental malperfusion than in singleton PreE cases (12, 13), implying that factors in addition to placentation deficiency may play a role in PreE in twins. Additionally, because uterine Doppler abnormalities indicative of uteroplacental insufficiency were found to be less common in PreE twin pregnancies with normal growth (14), and the future risk of maternal cardiovascular disease may be reduced in cases of PreE with twins compared to PreE with singletons (15), it is reasonable to propose that, compared to SGs, different pathophysiological pathways involving the fetoplacental and maternal systems work simultaneously to elicit the PreE syndrome in multifetal gestations.

Diamniotic twins may be dichorionic (with individual or fused placentas) or monochorionic (with a shared, single-disk placenta), while monoamniotic twins are invariably monochorionic. Dichorionic twinning is more common than monochorionicity, while monoamnionicity is infrequent (16). Clinical and epidemiological studies of PreE risk in association with chorionicity, however, have yielded inconsistent results (9, 17–19), possibly relating to multifarious factors that contribute to the complexity of such studies.

Considering the placental contribution to the pathophysiology of PreE (20), in this study we adopted a functional genomics approach to better understand the placental contribution to the pathobiology of PreE in twin gestations. We hypothesized that compared to singletons with PreE, twins with PreE would display changes in the placental transcriptome that align with established patterns of pathway dysregulation, such as placental hypoxia, altered metabolism, and angiogenic dysregulation. Prior to our study it was unclear whether, in dichorionic twin pregnancies, a molecular signature characteristic of PreE appeared in both placentas or only one at the time of clinical manifestation. Our goals were to: (1) ascertain the placental gene expression profiles and molecular pathways that distinguish PreE in twins from that in singletons; (2) correlate the placenta transcriptome with the clinical severity of the disease in twin gestations; and (3) estimate the degree of transcriptional concordance between twin placentas in relation to maternal disease status and chorionicity.

## Methods

Supporting data are available in the article and its Supplement. The sequencing data generated for this manuscript were deposited in the NCBI Gene Expression Omnibus (GEO) and are accessible through accession number GSE272342.

### Study Approval

This study was approved by the IRBs of Yale University, The Ohio State University, the Abigail Wexner Research Institute at Nationwide Children’s Hospital, and the University of Illinois at Chicago. All participants provided written informed consent.

### Recruitment and Tissue Collection

Transcriptomics analysis was performed on 32 individual villous placenta samples from 16 twin pregnancies classified by chorionicity (dichorionic, DT; and monochorionic, MT) and PreE diagnosis in the following groups: Group 1, normotensive singleton gestations (SGs) (n=10, gestational age at delivery [GA, mean±SD]: 35.3 ± 4.1 weeks);) from GSE203507 as previously reported (21); Group 2 No PreE DT (n=7, GA: 32.2 ± 3.4 weeks); Group3 No PreE MT (n=2, GA: 36.2 ± 0.1 weeks); Group 4 PreE DT (n=5, GA: 33.1 ± 4.7 weeks); Group 5 PreE MT (n=2, GA: 32.9 ± 3.7 weeks); Group 6 SGs across a spectrum of PreE clinical severity (n=10, GA: 34.0 ± 3.5 weeks); and Group 7 SG early-onset PreE cases (n=15, GA: 30.2 ± 2.8 weeks) from GSE203507. Clinical and demographic details of the included cases are presented in Table S1.

PreE was established clinically using the diagnostic criteria put forth by American College of Obstetricians and Gynecologists (ACOG) (22). For the purposes of this study, all PreE cases had hypertension >140/90 mmHg and proteinuria (>300 mg/24 h) after 20 weeks of gestation. Chorionicity in twin gestations was determined by antenatal ultrasound. Villous tissue biopsies from all placentas were obtained and processed immediately after delivery, as previously described (23). For each of the DT pregnancies, samples from the A and B twin placentas (as determined ultrasonographically) were collected individually; for the MT cases, two separate biopsies were obtained from each shared placenta (designated A1 and A2). To estimate actual fetal growth in relation to growth under typical physiological conditions, expected fetal weights were calculated as previously described (24). Deviation from the Hadlock weight (Δ_Hadlock_) was calculated by subtracting the birth weight from the Hadlock expected weight.

<u>Inclusion criteria</u>: Preeclampsia based on well recognized ACOG criteria, monochorionic and dichorionic implantation based on ultrasonograhic evaluation confirmed by pathological examination of the placenta, no associated medical co-morbidities (i.e, pre-gestational or gestational diabetes, chronic hypertension, connective tissue disorders). Absence of chronic co-morbidities was applicable to the singleton control placentas used in this study.

<u>Exclusion criteria</u>: Uncertain placentation, major associated medical co-morbidities, twin-to-twin transfusion syndrome or any other major placental perfusion imbalance between twins (i.e, twin reversed arterial perfusion [TRAP] sequence). MTs that received fetoscopic laser photocoagulation were excluded.

### RNA Extraction and Bulk RNA Sequencing (RNA-seq)

Total RNA was isolated from flash-frozen villous placental tissues using TRIzol reagent (Life Technologies, Carlsbad, CA) (23). For library preparation, the TruSeq Stranded Total RNA Sample Prep Kit with Ribo-Zero Gold (Illumina, San Diego, CA) was used, followed by RNA-seq using the Illumina HiSeq 2500 system to generate paired-end, 150 bp reads. Following a quality assessment with FastQC v0.11.8 (25), sequencing adapters were trimmed using Trimmomatic v0.39 software (26) and mapped to the hg38 genome assembly using TopHat2 v2.0.14 (27). Feature counts were generated with HTseq v0.11.2 (28).

Three early-onset PreE specimens from GSE203507 were re-extracted and sequenced *de novo* with the current sample group to assess variance attributable to changes in sequencing protocol. Three new samples from placentas used in our prior study (corresponding to S06 [GSM6175121], S18 [GSM6175133], and S20 [GSM6175135] from GSE203507) were extracted and prepared using the paired-end library and sequencing protocol to assess how the present methodology compared to the original protocol (single-end, 50 bp reads) (21). The singleton PreE placentas were sequenced interspaced among the twin placentas to mitigate further technical bias.

### Integration with Previously Published Datasets

To establish a benchmark cohort of mostly SG samples with early onset PreE and controls (No PreE), sequencing and phenotypic (clinical) information were abstracted from three prior RNA-seq studies involving villous placental tissue samples: (1) GSE114691 (29), comprising 61 samples (40 with PreE, and 21 from normotensive preterm births used as controls); (2) GSE148241 (30), consisting of 41 samples (9 with PreE, 32 term controls); and (3) GSE203507 (21), contributing 25 samples (15 PreE specimens, 5 preterm controls, and 5 term controls). The last of these was from our prior report on placental transcriptomics in early-onset PreE. These 127 specimens established the 3-cohort benchmark PreE dataset, details of which are as summarized previously (21).

The same analytical workflow described above was used to generate feature counts from the available raw FASTQ files, with application of single- and paired-end trimming and mapping protocols depending on library type. Presumed fetal sex was estimated based on the placental expression of selected sexually dimorphic transcripts (*XIST*, *TTTY15*, *TXLNGY*, *EIF1AY*, and *ZFY*). Where available, these designations were confirmed using associated study data. For combined analyses, the datasets were adjusted for fetal sex and cohort-dependent batch effects, as described below.

### Data Transformations, Batch Correction, and Differential Abundance Analysis

The goodSamplesGenes function from the WGCNA v1.72-1 (31) R library was used during preprocessing to identify missing entries and zero-variance genes. Of the original 26,485 expressed genes in the dataset, 1,213 transcripts were removed in this process.

Variance stabilizing transformed (vst) expression data were generated using the vst function from DESeq2 v1.37.5 (32), and the results were corrected for cohort and fetal sex using the removeBatchEffect function from the limma v3.53.5 R package (33). The effects of the data adjustments on principal component analysis are shown in Fig. S1 for the samples from the present study and from the 3 prior datasets. The three new samples from the same original placentas used in the GSE203507 study showed high degrees of correlation in the final batch-corrected data: S06 (r=0.995, ρ=0.969); S18 (r=0.992, ρ=0.959); and S20 (r=0.994, ρ=0.967). The sample designations were as we reported previously and were all from severe early-onset PreE cases (21).

Statistical testing for differential transcript abundance was performed with DESeq2 (32). For the 3-cohort benchmark dataset, the multi-factor design formula included the presence or absence of PreE, the cohort from which the samples originated, and fetal sex. For the data from the twins cohort, including the 32 individual twin samples, the newly sequenced singleton placentas (10 PreE), and reference samples from GSE203507 (15 PreE specimens, 10 controls), the raw counts were adjusted for library type using the ComBat_seq function from sva (Surrogate Variable Analysis) v3.48.0 (34) then analyzed by DESeq2 with assigned fetal sex as a covariable in the multi-factor design formula. To evaluate the robustness of the twins cohort models, we performed a sensitivity analysis using subsets of the full model grouped by sequencing library type (single- or paired-end). The significance of coefficients in the resulting generalized linear models was evaluated using Wald (nbinomWaldTest function) and likelihood-ratio (nbinomLRT function) significance testing. The threshold for statistical significance was false discovery rate (FDR)<0.05 using the Benjamini-Hochberg procedure (35), unless indicated otherwise.

### Derivation of the PreE Molecular Classification Signature

A machine learning approach implemented with DaMiRseq v2.9.0 (36) was used to generate a small set of non-redundant transcripts capable of reproducibly partitioning a majority of PreE placenta samples from controls. For this analysis, we used the 3-cohort benchmark dataset comprising normotensive and early-onset severe PreE placental samples (n=127). Briefly, the DaMiRseq normalization procedure was used to generate vst counts, which were then adjusted for 4 surrogate variables to correct for confounding factors. Next, feature selection for binary classification (PreE vs. No PreE) was performed using class-correlated principal components and a partial least squares backward variable elimination procedure over 10 iterations (the algorithm takes the intersection among individual feature sets to enrich for robust features). Finally, highly correlated (|ρ|>0.85) features were excluded from the set of informative transcripts. Feature importance was calculated using the regressional ReliefF algorithm (37).

To evaluate the performance of the final signature for binary classification, we employed the DaMiR.EnsembleLearning function. From the 3-cohort dataset, bootstrap sampling (70/30 training/testing) was used to generate training and test samples consisting of either the molecular classifier transcripts or an equivalent number of randomly selected transcripts. The training set was sampled to generate nested sets to train and cross-validate 5 statistical learning classification algorithms (random forest, k-nearest neighbors, linear discriminant analysis, naïve Bayes, and support vector machine). From these, an ensemble classifier was generated through a weighted majority voting approach involving a linear combination of the weights and predictions of the individual classifiers.

Classification performance was then assessed using the original data held over for testing. The resampling process was repeated over 100 iterations.

### Sample Distance Metrics

<u>Distance along PreE spectrum</u>. We estimated the distance of each sample along a spectrum of placental transcript expression ranging from that characteristic of uncomplicated gestations (No PreE) to that observed with severe, early-onset PreE using the 98-transcript PreE molecular classification signature. This signature was first evaluated using a rank-based, single-sample scoring algorithm implemented with singscore v1.17.0 (38). The Singscore expression score (ES) was obtained by scoring individual samples based on the 98-transcript molecular classifier (32 transcripts decreased with PreE, 66 increased with PreE). First, the full set of 25,272 transcripts was ranked using the rankGenes function; this was followed by application of the simpleScore function to the ranked expression data using the bidirectional target classification signature described above, and using the resulting total score as the final ES. High-scoring samples were interpreted as having transcriptomes most concordant with the PreE classification signature, with individual scores reflecting the relative mean percentile rank of the target signature within each sample.

As a complementary approach, we also calculated the Mahalanobis distance between individual samples and the centroid for non-PreE placental specimens in principal component space (top two coordinates, PC_12_) after applying the 98-transcript molecular classifier to batch-corrected vst counts. The mahalanobis function in the R stats package was used for this.

<u>Distance between paired samples</u>. We generated two distance metrics to assess the degree of dissimilarity between dichorionic sibling twin pairs. First, we calculated a Poisson dissimilarity matrix using the PoiClaClu v1.0.2.1 R package (39) based on the batch-corrected vst expression for the 98-transcript classifier. We calculated Spearman correlation distance (1 – ρ) for the same 98 transcripts as a secondary assessment. These values were compared to the sample distances from the MT (two independent samples from the same placenta) and the three SG samples (S06, S18, and S20) that were extracted and sequenced in both the present study and the GSE203507 dataset (21).

### Functional Enrichment Analyses

To identify the most dysregulated placental pathways in the setting of PreE, we first performed a global functional enrichment analysis for the 3-cohort benchmark dataset. Overrepresentation analysis (hypergeometric test) was evaluated using the enricher function in the clusterProfiler v4.5.2 R package (40). Specifically, we tested for enrichment of curated KEGG and WikiPathway gene sets obtained from the Molecular Signatures Database (MSigDB, v7.5.1) C2 sub-collection and combined (898 gene sets in total). A subset of differentially abundant transcripts in the PreE vs. No PreE comparison in the 3-cohort dataset (3,241 expressed genes, FDR<0.0001) were evaluated against the background of the remaining mapped transcripts; parameters included limiting gene sets to between 10 and 250 members in size, a threshold *p*-value<0.05, and FDR<0.05. Further annotation was accomplished using the AnnotationDbi v1.59.1 package and the Bioconductor annotation package org.Hs.eg.db v3.14.0. Additional scoring was performed using the get_aggrscores function from the GeneTonic v2.1.3 R library (41).

Next, to evaluate the variation of pathway activity among individual samples, we calculated a non-parametric gene set variation analysis (GSVA) score using sample-wise gene set variation analysis implemented with the GSVA v1.45.2 R package (42). For this, the 3,241-transcript signature used in the global PreE overrepresentation analysis was applied to the above KEGG and WikiPathway gene sets having between 10 and 250 members. GSVA scores were prioritized by applying Mann-Whitney (for two-group comparisons) or Kruskal-Wallis testing (for comparisons of three or more groups) with FDR multiple comparisons adjustment using the Benjamini and Hochberg procedure.

### Metabolic Reaction Activity Scoring

To infer the relative metabolic activity in villous placental tissue from the gene expression data, we used METAFlux v0.0.0.9 (43) to calculate sample-specific metabolic reaction activity (MRA) scores for each of the 13,082 reactions cataloged in the Human1 genome-scale metabolic model (44). MRA scores were determined from batch-corrected vst expression data (25,289 expressed transcripts) using the METAFlux calculate_reaction_score function and evaluated using Mann-Whitney or Kruskal-Wallis testing as appropriate, with FDR adjustment of *p*-values. Spearman correlations evaluated the relationships between the MRA scores and GSVA pathway scores.

### Data Visualization

The ggplot2 v3.3.6 R library was used for data visualization, with extensions including the GGally, ggExtra, gridExtra, patchwork, ggMarginal, ggpubr, and ComplexUpset packages. Other functions used or adapted from the R libraries were noted in the figure legends. Heat maps were produced using pheatmap v1.0.12. Additional plots and alternate renderings were generated using Prism v10.1.2 (GraphPad Software, La Jolla, CA).

### Statistics

Statistical analyses were performed using multiple functions and packages in R (v4.2.1 or greater) and Bioconductor (v3.15 or greater). Additional statistical tests were conducted using Prism v10.1.2 (GraphPad Software, La Jolla, CA). In most cases, non-parametric statistical methods were favored over parametric methods, although in all instances, normality was first tested using the Shapiro–Wilk test. The table summary of clinicodemographic characteristics was aided by the use of the sTabl3R R package v0.3 (45). Principal component analysis was achieved using the prcomp function in the base R v4.2.1 stats library or the plotPCA function in the DESeq2 package, as indicated in the figure legends. Univariate and multivariate linear regression models were fitted using the lm function of R stats (for fixed effects), and lme4 v1.1-35.3 with lmerTest v3.1-3 (for mixed effects).

Generalized estimating equation (GEE) models (used to account for correlation between twins) were implemented with the glmgee function in glmtoolbox v0.1.11. Correlation analyses (Pearson and Spearman) were performed using either cor.test in R stats or rcorr from the Hmisc v5.1-1 R library. Point-biserial correlations (to measure the degree of association between continuous variables and dichotomous factors) were performed using the ltm (Latent Trait Models) v1.2 R package. Additional statistical tests used were as indicated in the preceding methodological subsections. Unless otherwise stated, a two-sided *p*-value<0.05 was considered statistically significant.

## Results

### Overview of Study Population and Differential Abundance Analysis

Transcriptional profiling by RNA-seq was performed on 32 individual villous placental specimens from 16 carefully phenotyped twin gestations grouped by maternal disease status (normotensive, n=9; or PreE, n=7) and chorionicity (dichorionic twin, DT, n=12; and monochorionic twin, MT, n=4): We studied the following groups: **Group 1**, the referent group, comprised normotensive singleton gestations (SGs) (Group 1, n=10 samples) from GSE203507 as we previously reported (21). **Groups 2** and **3** comprised DT and MT placentas without PreE (n=7 and n=2 pairs, respectively), while **Groups 4** and **5** consisted of DT and MT placentas with PreE (n=5 and n=2 pairs, respectively). **Group 6** included 10 samples from SGs across a spectrum of PreE clinical severity. The remaining 15 samples were SG early-onset PreE cases (**Group 7**; n=15 samples) from GSE203507. This group was included to establish a boundary position along the continuum against which the twin placentas were evaluated. The clinical groups summaries and detailed characteristics of the individual samples are presented in Table S1.

Principal component (PC) analysis revealed batch effects between the samples of the present study and those of our prior analyses (GSE203507 samples in Fig. S1A). Therefore, for differential abundance analysis, in addition to the clinical group, the full statistical models included adjustments for batch effects and differences attributable to assigned fetal sex as reflected in the placental transcriptome. By the likelihood-ratio test across all 7 groups (relative to a baseline model in which the clinical group coefficient was omitted), there were 2,946 variably abundant transcripts (FDR<0.05).

Based on these transcripts, we observed that the DT normotensive placentas (Group 2) most closely resembled the normotensive SG specimens (Group 1), while the PreE DT (Group 4) were most like the SG PreE samples (Groups 6&7) (Fig. 1A-C). The PreE MT (Group 5) exhibited a graded similarity pattern relative to the other groups, resembling that of the PreE DT (Group 4) (Fig. 1A). Interestingly, we observed a large heterogeneity among MT cases. Analysis of the individual samples revealed similarity score co-clustering of 2 pairs of No PreE MT and PreE MT samples with the SG Group 6&7 PreE samples (Fig. 1B), which was less prominent in the clustering based on transcript abundance (Fig. 1C).

**Figure 1.**
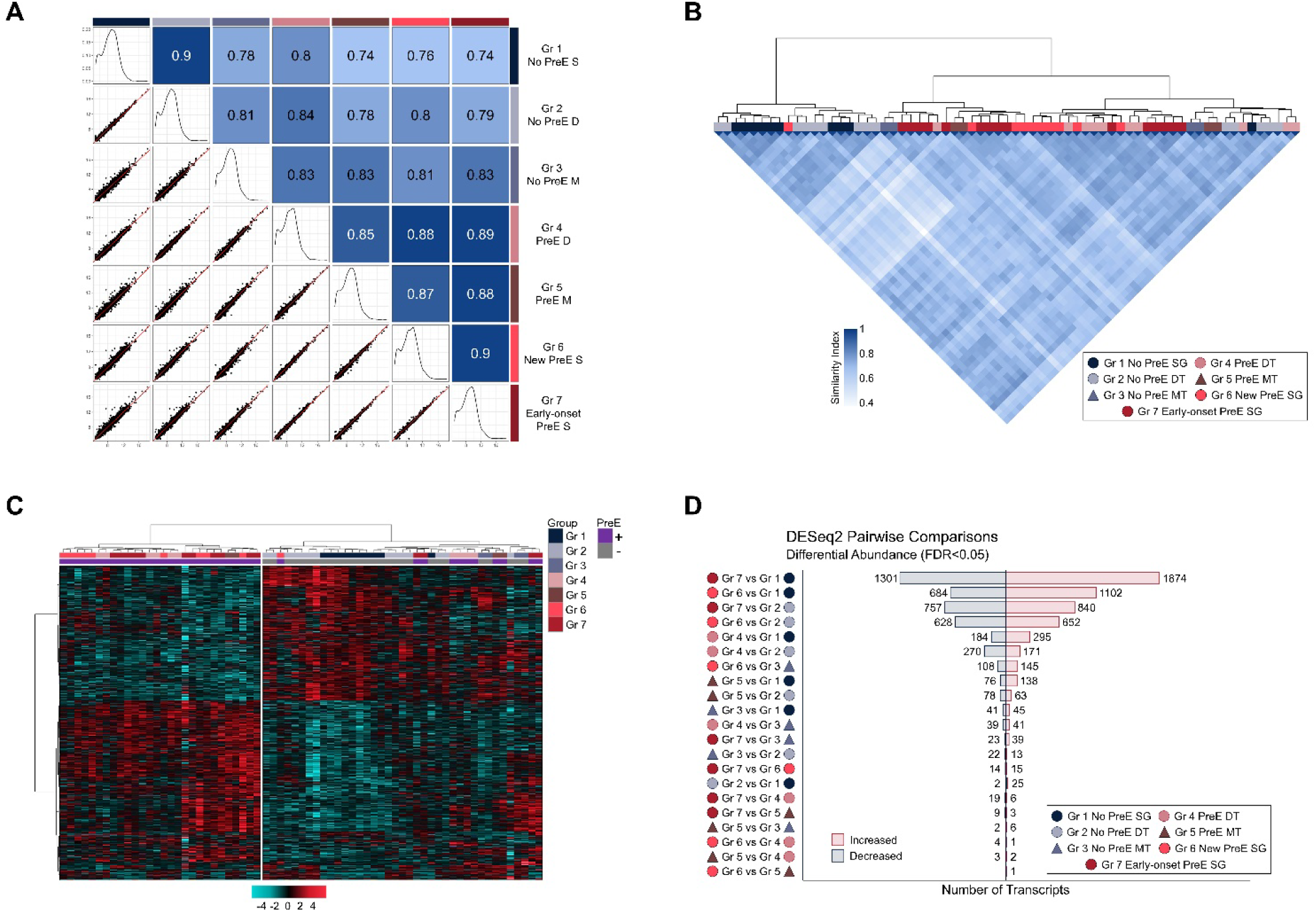
Differential abundance analysis for twins cohort. RNA-seq was performed on a twins cohort that included 32 individual twin samples, 10 newly sequenced singleton placentas, and 25 reference samples from GSE203507 (15 early-onset PreE specimens, 10 controls). Differential abundance across clinical groups was determined with DESeq2 by the likelihood-ratio test (LRT) on models of batch-corrected counts; the full model included both clinical group and assigned fetal sex as coefficients, while the reduced model included only assigned fetal sex. Pairwise differential abundance was calculated with DESeq2 using the full model with Wald significance testing for between-group comparisons. (A) Pairs plot showing scatterplots (lower triangle), density plots (diagonal), and the similarity matrix (upper triangle) for pairs of clinically defined groups. Values in scatterplots (lower triangle) represent the mean expression of 2,946 (FDR<0.05, LRT) batch-corrected, variance-stabilizing transformed transcripts per group. Similarity values (upper triangle) were based on the mean expression of these same transcripts and calculated as the complement of the scaled Euclidean distance. The scale is arranged by color saturation from light (most dissimilar) to dark (identity). (B) Heat map with unsupervised hierarchical clustering showing similarity values for the individual samples with the color scale as in panel A. (C) Clustered heat map demonstrating relative expression of the 2,946 differentially abundant transcripts (FDR<0.05, LRT). (D) Stacked bar plot showing the results for pairwise group comparisons from DESeq2 full statistical models (FDR<0.05, Wald test). Abbreviations: DT, dichorionic twin; Gr, group; LRT, likelihood-ratio test; MT, monochorionic twin; PreE, preeclampsia; SG, singleton gestation.

For the subsequent 21 pairwise group comparisons, we used the full DESeq2 model described above with Wald significance testing between groups. These results generally mirrored that of the similarity analysis in that the group pairs with the greatest divergence (Group 7 [SG early-onset PreE cases] vs Group 1 [SGs normotensive singleton gestations]) tended to produce the most variably abundant transcripts, while the most similar group pairs (Group 6 [SGs across a spectrum of PreE clinical severity] vs Group 5 [MT placentas with PreE]) yielded the fewest (Fig. 1D). Exceptions to this general trend were observed: the 2 group pairs with the highest similarity scores (Groups 6&7 [SGs across a spectrum of PreE clinical severity and SGs with early onset-PreE] and Groups 1&2 [normotensive SGs and DT without PreE], both 0.9, Fig. 1A) had larger expression differences (29 and 27, respectively) than were observed in an additional 6 pairwise group comparisons (Fig. 1D). Such discrepancies, particularly for the MT groups, indicated that differential abundance analyses alone would be insufficient to stratify the twin placentas.

To evaluate the robustness of the pairwise results, we created additional statistical models for each of the two batches individually (Fig. S2). Despite not allowing a full set of pairwise comparisons, these batch-specific models demonstrated similar differential abundance magnitudes compared to the full model, with considerable transcript overlap for synonymous pairwise contrasts at a nominal cut-off of FDR<0.05 (Fig. S2). The results of both models are presented in Tables S2&S3.

### Development of a Molecular Classifier for Quantification Along a Placental PreE Spectrum

We next sought to integrate and organize the placental transcriptional profiles along a quantitative spectrum. To generate a consensus signature to facilitate PreE classification, we used an integrated RNA-seq dataset comprising normotensive control samples and specimens with early-onset PreE from 3 independent cohorts of human villous placental tissue (127 samples from datasets GSE114691, GSE148241, and GSE203507) (21, 29, 30). Starting with the full transcriptional profiles, we applied binary classification with backward feature elimination using DaMiRSeq software (36).

From 25,272 mapped transcript counts, 11,301 features were filtered out based on expression criteria (a minimum of 10 feature counts in at least 70% of samples). Of the 13,966 features remaining, we determined that 192 candidates were selected as being the most informative for classification.

Following the removal of highly correlated features, a final list of 98 transcripts established the final PreE molecular classification signature (Table S4, Fig. S3). The final classifier contained placental transcripts previously identified as having increased expression in PreE, including *AQP1*, *BCL6*, *CGA*, *CRH*, *EBI3*, *INHA*, *PHYHIP*, *SIGLEC6*, *SLCO2A1*, *SPAG4*, and *TREM1* (46). We further noted that, prior to the removal of highly correlated features, the penultimate list included many additional PreE-associated transcripts such as *ENG*, *FLT1*, *HEXB*, *HTRA4,* and *NRIP1* (46–49). PC analysis using the molecular classifier resulted in increased separation between the control (No PreE) and PreE clusters (Fig. 2C) compared with the full set of transcripts (Fig. 2A). Furthermore, for binary classification (PreE vs. No PreE) of the 3-cohort benchmark data (n=127 samples), the 98-transcript signature (Fig. 2B) outperformed a panel of 98 randomly selected transcripts across a variety of supervised learning algorithms (Fig. S3).

**Figure 2.**
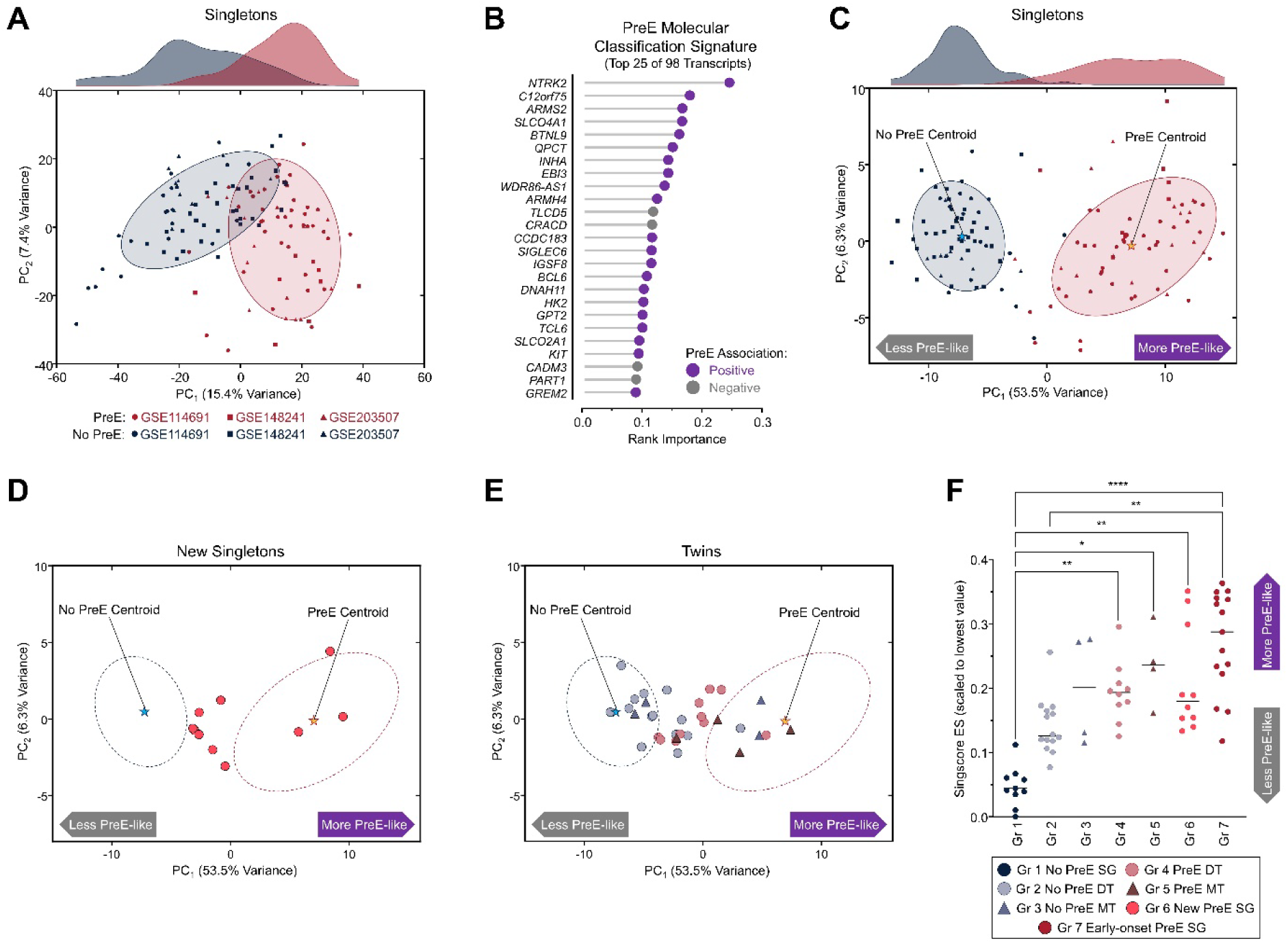
Establishment and application of 98-transcript classifier for molecular stratification of villous placental samples. DaMiRseq was used to identify a small set of consensus transcriptional biomarkers and for classifying the 3-cohort benchmark early-onset PreE dataset (n=127 samples from GSE148241, GSE114691, and GSE2035070). Feature selection resulted in a panel of 98 transcripts reliably segregating most samples with early-onset preeclampsia (PreE) from controls (No PreE). (A) Principal component (PC) analysis for the global transcriptome (25,272 transcripts) of the 3-cohort dataset. (B) Dot plot showing the top 25 of 98 transcripts identified in the classification signature, ranked by regressional ReliefF. Transcripts positively associated with PreE are represented in purple, while negatively associated features are shown in gray. (C) PC_12_ for the 3-cohort dataset using the PreE molecular classifier (98 transcripts). Stars indicate centroids; ellipses indicate the 80% confidence levels for a multivariate t-distribution for each condition. PC_1_ accounted for 53.5% of the variance in the dataset and was strongly associated with the absence or presence of early-onset PreE (point-biserial correlation 0.90, U statistic 2049, *p*<0.001, Mann-Whitney test) but not with fetal sex (point-biserial correlation −0.05, U statistic 1881, *p*=0.52, Mann-Whitney test). (D) Projection of new SG samples onto PC_12_ based on PreE molecular classifier expression. Dotted ellipses show 80% confidence levels for the PreE and control (No PreE) samples from the full 3-cohort dataset. (E) Distribution of twin placenta samples within PC_12_. Dotted ellipses are as in panel D. (F) Plot of Singscore ES (scaled to the lowest calculated value) showing individual values for each group, with bars depicting medians. High-scoring samples represent expression signatures that were more consistent with PreE. Group medians were evaluated using the Kruskal-Wallis test (H statistic 39.83, *p*<0.0001). Asterisks indicate significance levels in multiple comparisons using the Dunn test: *, *p*<0.05; **, *p<*0.01*; ****, p*<0.0001. See also Figure S7 for positions of individual twin pairs. Abbreviations: DT, dichorionic twin; ES, expression score; Gr, group; MT, monochorionic twin; PC, principal component; PreE, preeclampsia; SG, singleton gestation.

Encouraged by these results, we proceed to evaluate whether this expression signature could be used to stratify specimens along a continuous range. We began by considering PC analysis for the newly sequenced singleton samples (PreE, n=10). The introduction of these samples produced a distribution that ranged from between the 80% CI of the No PreE centroid to within that of the PreE centroid, with most (7/10) of the Group 6 samples being distributed in the intermediate range along PC_1_ (Fig. 2D).

Having established the utility of the classifier in the singleton cases, we proceeded to apply the molecular classifier to placentas from twin gestations. In the PC_12_ coordinates, 8/14 individual DT No PreE (Group 2) samples were within the 80% CI of the No PreE centroid, and all but one of the remainder fell outside of the PreE centroid 80% CI (Fig. 2E). For the DT PreE (Group 4) specimens, 9/10 of the individual samples were clustered between the centroid CIs. Curiously, proximity to the PreE centroid was dominated by MT samples.

To better quantify and compare the relationships among the groups along the PreE spectrum, we applied the PreE classification signature to score individual samples using two distance metrics: (1) the Singscore ES metric, a non-parametric rank-based expression scoring algorithm (38); and (2) the Mahalanobis distance from 3-cohort No PreE centroid, which provided a measurable estimate of the observed distributions in PC_12_ space. We found that the Singscore ES was highly correlated with Mahalanobis distance (r=0.92, *p*<0.0001; ρ=0.97, *p*<0.0001), indicating that these estimates were robust (Fig. S4). Both the Singscore ES and Mahalanobis distance metrics were strongly correlated with placental *FLT1*, *ENG*, and *LEP* mRNA expression, as well as the ratio *FLT1*/*PGF* (Fig. S5).

Correlation was anticipated for *FLT1* and *ENG* since these were omitted from the classification signature due to redundancy; nonetheless, the degree of these correlations was striking (r≥0.8 in combined cohort) and strengthened our confidence in the validity of the scoring systems.

Each of the transcript-based placental scoring metrics was positively correlated with the highest recorded maternal blood pressure and negatively correlated with birthweight (Fig. S6). Furthermore, the Singscore ES and Mahalanobis distance metrics were significantly correlated with the reduction in birthweight from the Hadlock expected fetal weight (Δ_Hadlock_), and regression modeling demonstrated that both were independent predictors of suboptimal fetal growth as estimated by this deviation (Fig. S6, Table S5).

Since the Singscore ES exhibited an expanded linear range compared to the Mahalanobis distance metric (Fig. S4C), we prioritized the former in our subsequent analysis. Significant differences in Singscore ES were observed among the 7 groups (H statistic 39.83, *p*<0.0001, Kruskal-Wallis test), with medians of all 4 of the PreE singleton and twin (DT and MT) groups (Groups 4-7) being greater than that of the control samples (Group 1, SG No PreE) (Fig. 2F; see also Fig. S7 for positions of the individual twin pairs). Interestingly, the DT No PreE samples (Group 2) also had a greater median Singscore ES than the SG controls (Group 1). Following multiplicity adjustment, however, this did not reach a level of statistical significance (uncorrected Dunn *p*=0.036, multiplicity adjusted *p*=0.148).

### Divergence in the Placental PreE Classification Signature Between Sibling Twin Pairs

Next, we applied the molecular classifier to assess whether placental transcriptome dissimilarities between sibling placenta pairs characterized PreE in DT pregnancies. As a reference, we also calculated the distances between the singleton placental samples (all from cases with severe early-onset PreE) that were extracted and sequenced in both the present study and the prior GSE203507 dataset; these placentas provided an estimate of the expected distance for the MTs (in which two samples of each shared placenta were sequenced). Transcriptional discordance was determined using both Poisson dissimilarity and Spearman correlation distance; these distance metrics were strongly correlated (r=0.97, *p*<0.0001; ρ=0.95, *p*<0.0001) (Fig. S8). Overall, the MT samples (Groups 3&5) were the most similar among the sibling placenta pairs (Fig. 3, Fig. S8), having distance metrics similar to those for the reference placentas; the MT PreE pairs were slightly more discordant than the MT control pairs. Both groups of DT pairs (Groups 2&4) exhibited a broader range of distance values: The median [range] Poisson dissimilarity for the DT No PreE (Group 2) samples was 12.1 [7.8-31.4], while that for the DT PreE (Group 4) samples was 13.2 [12.4-32.9] (Fig. 3), and the corresponding Spearman correlation distances were similar (Fig. S8). By either metric, the median dissimilarity for the DT PreE (Group 4), but not the DT controls (Group 2), was significantly greater than that of the reference (Fig. 3, Fig. S8).

**Figure 3.**
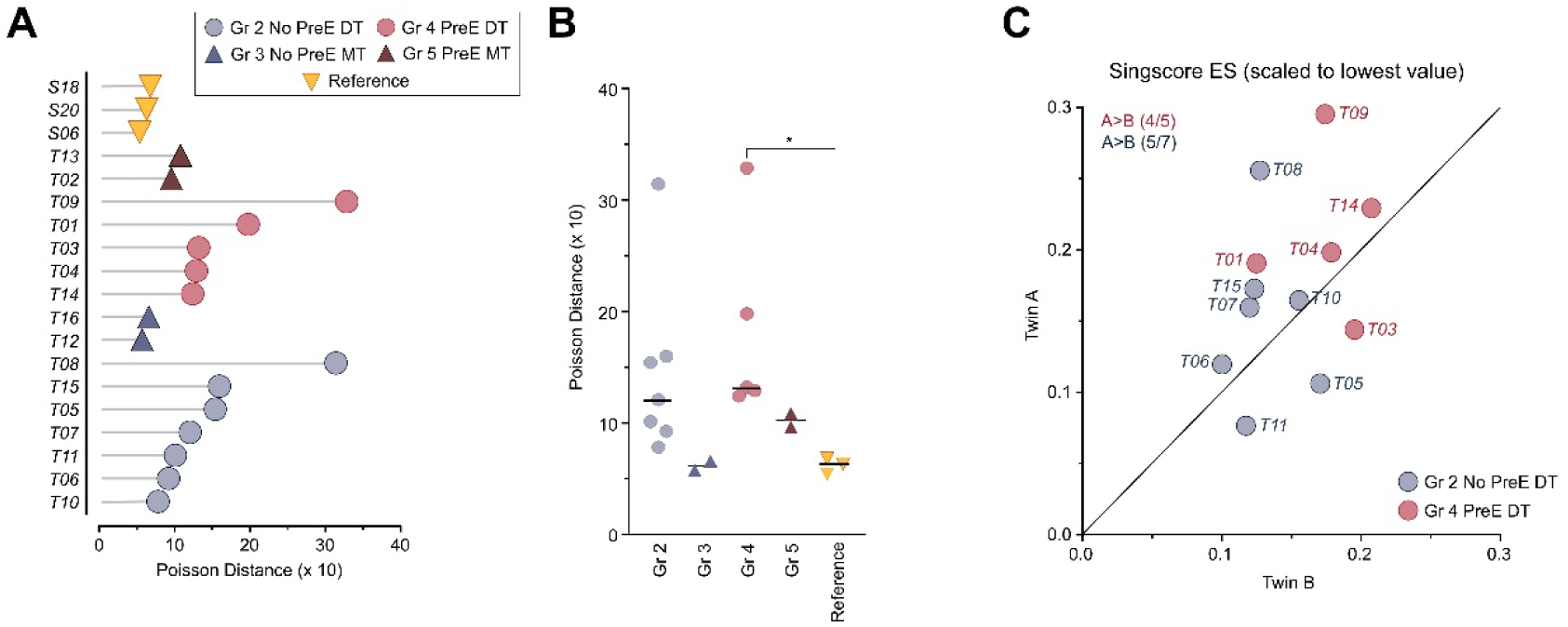
Evaluation of between-sample transcriptomic distances for sibling twin placentas. Using the 98-transcript PreE molecular classification signature, we calculated distances between twin sibling placental sample pairs based on Poisson dissimilarity. As a reference, we calculated the distances between the SG placental samples (all from cases with severe early-onset PreE) extracted and sequenced both in the present study and the prior GSE203507 dataset. (A) Dot plot of between-sample Poisson distance for the twin and reference samples. Increasing distance indicates less similarity. (B) Plot of Poisson distance showing individual values for each group, with bars representing medians. Medians were evaluated using the Kruskal-Wallis test (H statistic 12.4, *p*=0.015). Asterisk indicates *p<*0.05 (Dunn test). (C) Scatterplot of Singscore ES as calculated in Fig. 1F for Twin A versus Twin B in DT placentas. A higher ES score indicates a transcriptome that is more consistent with PreE. Abbreviations: DT, dichorionic twin; ES, expression score; Gr, group; MT, monochorionic twin; PreE, preeclampsia.

Among DT placentas, that from the A (presenting) twin had a greater Singscore ES (i.e., a transcriptional profile more consistent with a PreE-like expression signature) than the B twin in 9/12 cases (No PreE: 5/7; PreE: 4/5; *p*=0.0730, binomial test).

### Twin Placentas Show Heterogeneity in PreE-Associated Functional Dysregulation

To identify consensus dysregulated pathways in placenta specimens in the setting of PreE, we first performed a global functional enrichment analysis for the 3-cohort benchmark dataset. Using the top 3,241 transcripts from the PreE vs. No PreE comparison (FDR<0.0001, Wald test), this analysis established 12 statistically significant consensus gene sets for further evaluation (FDR<0.05, hypergeometric test) (Fig. 4). Focusing on these pathways, we next applied individual sample scoring (gene set variation analysis, GSVA) to the samples from our prior dataset (GSE203507) and the 42 newly sequenced samples.

**Figure 4.**
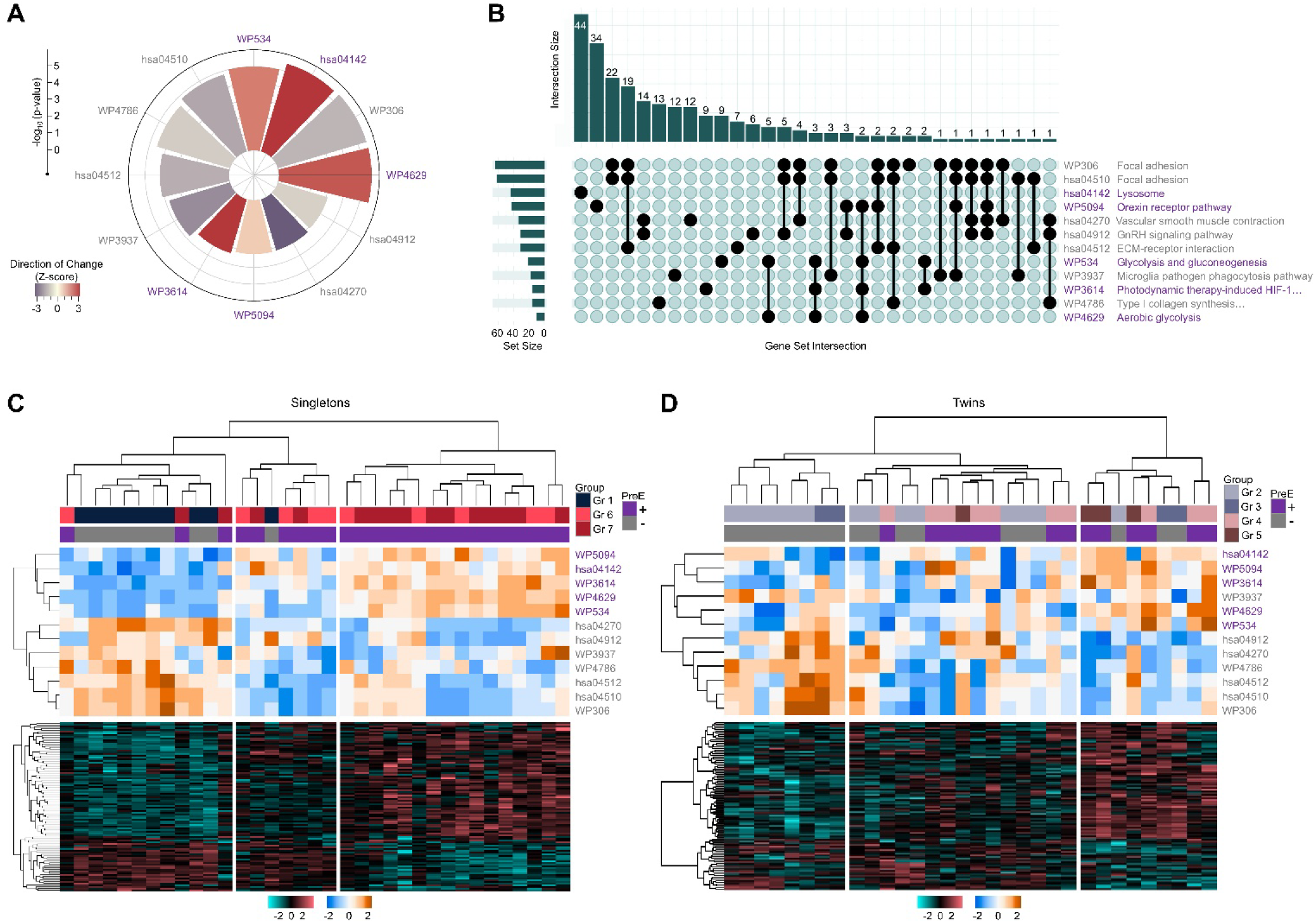
Functional enrichment analysis for individual samples. To identify consensus dysregulated pathways in placenta specimens associated with PreE, we first performed a global functional enrichment analysis for the 3-cohort benchmark dataset (n=127 samples). Using the top 3,241 transcripts from the PreE vs. No PreE comparison (FDR<0.0001, Wald test), 12 gene sets were significantly enriched (FDR<0.05, hypergeometric test). Sample-level gene set variation analysis (GSVA) sores for these pathways were calculated using the GSVA package. (A) Circular barplot showing the top 12 most overrepresented pathways in the global functional enrichment analysis, with bar height representing −log_10_ (*p*-value) and color representing the Z-score (an estimate of the direction of the change, with positive scores indicating that most of the gene set transcripts showed an increase in abundance in the setting of PreE). Purple and gray font colors indicate gene sets having positive and negative Z-scores, respectively. (B) UpSet plot showing sizes of gene sets and their intersections (genes in common) for the top 12 pathways most overrepresented in the 3-cohort dataset. (C) Clustered heat map of GSVA scores for individual SG placental samples from the current study with GSE203507 samples as reference (top of panel), and a corresponding heat map showing relative expression of the 98-transcript PreE molecular classifier (bottom of panel). (D) Clustered heat map of GSVA scores and expression of PreE molecular classifier transcripts as in panel C, but for the twin placental samples. Abbreviations: DT, dichorionic twin; Gr, group; GSVA, gene variation analysis; MT, monochorionic twin; PreE, preeclampsia; SG, singleton gestation.

For the SG placentas, unsupervised clustering of the GSVA scores revealed 3 general patterns:

(1) a set comprised mostly of No PreE (Group 1) samples; (2) a smaller, intermediate set; and (3) a large set of PreE specimens (Fig. 4C). For the twin samples, although cluster analysis also partitioned the individual samples into 3 similarly arranged groups, it was the intermediate cluster that contained the largest proportion of intermixed PreE and No PreE samples (15/32) of samples (Fig. 4D). This distribution recapitulated the arrangement observed by PC analysis (Fig. 2E).

To corroborate these results, we proceeded to evaluate the clustering of metabolic reaction activity (MRA) scores calculated using the METAFlux R package (43). This method marshals a genome-scale human metabolic network model to interrogate reaction activity based on the associated expression of metabolic transcripts. As an initial evaluation, we first applied MRA scoring to the 3-cohort dataset, which revealed 67 metabolic pathways (Fig. S9A) differentiating the PreE from the No PreE samples (FDR<0.05, Mann-Whitney test). There were moderate to strong correlations between the MRA and GSVA scores for several of the curated KEGG and WikiPathway gene sets. However, these relationships exhibited global heterogeneity (Fig. S9B-E), suggesting that the MRA scoring approach yielded information complementary to the GSVA analysis.

When MRA scoring was applied to the twins cohort dataset, 30 metabolic pathways differed across the 7 groups (FDR<0.05, Krusal-Wallis test) (Fig. 5A). For the SG placentas, MRA score cluster analysis yielded a PreE group dominated by Group 7 samples with high metabolic scores, a PreE group with intermediate metabolic scores consisting primarily of Group 6 samples, and a set with low metabolic scores comprising most Group 1 specimens (Fig. 5B). For the twin samples, a majority (21/32) had intermediate MRA scores, with only 5/32 individual PreE specimens exhibiting the high MRA scores characteristic of the Group 7 SG PreE samples (Fig. 5C).

**Figure 5.**
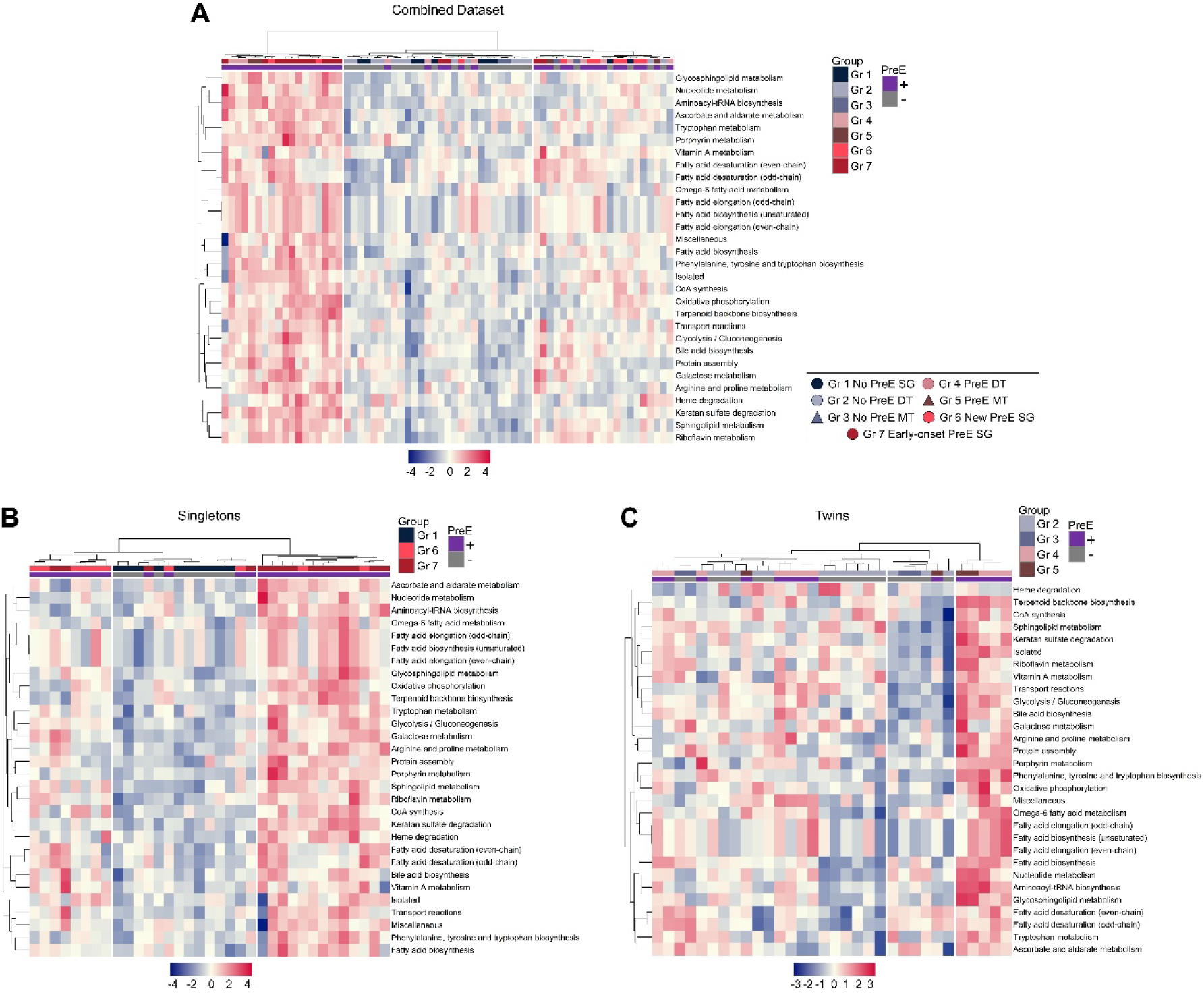
Metabolic reaction activity (MRA) scoring for individual samples in the twins cohort dataset. Pathway-level MRA scores were calculated with METAFlux software for 13,082 cataloged reactions using the batch-corrected, variance-stabilizing transformed expression data (25,289 transcripts) from the twins cohort dataset (n=67 samples). (A) Clustered heat map of the 30 pathway-level metabolic MRA scores that differed significantly across the 7 clinical groups in the cohort (FDR<0.05, Kruskal-Wallis test with *p*-value adjustment). (B) Clustered heat map of the MRA scores in panel A for the individual SG placental samples. (C) Clustered heat map of MRA scores in panel A for the individual twin placental samples. Abbreviations: DT, dichorionic twin; Gr, group; MT, monochorionic twin; MRA, metabolic reaction activity; PreE, preeclampsia; SG, singleton gestation.

Cluster analysis applied to each functional scoring procedure individually showed that most of the individual twin PreE samples (8/14 for GSVA, 9/14 for MRA scoring) were grouped separately from those with patterns more characteristic of SG PreE (Figs. 4D&5C). However, when considering the data collectively, we noted that most (11/12) of the individual samples displaying any PreE-like pathway dysregulation were also among the top third of all twin placentas when ranked by decreasing Singscore ES. Thus, despite the variability in response, those samples with more PreE-like expression signatures also exhibited more PreE-like functional perturbations. Interestingly, but also consistent with the molecular classifier scoring, the pathway clustering yielded an intermixing of twin samples of differing clinical PreE status. In summary, some twin samples without clinical PreE (at least at the time of delivery) possessed PreE-like transcriptional phenotypes.

## Discussion

In multifetal gestations, PreE has long been attributed, at least in part, to an overall increase in placental mass. In support of this hypothesis, hypertensive complications grow in proportion to the total number of fetuses (and fetoplacental units) present (50), whereas multifetal pregnancy reduction reduces PreE risk (13, 51). Indeed, the excess placental mass present in twin pregnancies, relative to singletons, contributes to elevated production of circulating factors (sFlt-1 levels and sFlt-1/PlGF ratios) that raise PreE risk (11, 52, 53). Although anti-angiogenic factor excess is an important determinant of maternal PreE symptoms, increased placental mass alone appears insufficient to elicit the clinical syndrome, as evidenced by the fact that most twin pregnancies are not complicated by hypertensive disorders (50).

Compelling data suggests that in twins, defective placentation and an increased immunologic response and/or semi-allograft incompatibility are central to PreE pathogenesis (8, 54). The clinical and epidemiological observations that DT gestations are frequently associated with discordant fetal growth implies that the placenta of the growth restricted fetus is hypoxic and thus defective placentation of just one placenta may trigger PreE in multifetal pregnancies (55). Alternatively, if the twin placentas were to exhibit the same PreE molecular phenotype, this might suggest that a shared systemic circulatory factor affecting both placentas could be responsible for the development of this hypertensive disorder. Interestingly, current epidemiological evidence does not support the expected increase in rates of hypertensive disorders among dizygotic versus monozygotic twins (8), nor is there consistent evidence that DTs have greater PreE risk than MTs (9, 17–19). Given the growing consensus that the pathogenesis of PreE in singletons is complex and multifactorial (5, 56), it is likely that PreE in multifetal gestations is just as intricate, if not more so.

This study examined the placental transcriptome and associated molecular pathways characterizing PreE in twin gestations. In addition to performing standard differential abundance analysis, we developed an expression signature to score individual placentas along a molecular spectrum of disease severity. Using a machine learning approach, we developed a 98-transcript classifier and determined that this accurately differentiated (normotensive placentas and placentas delivered preterm with severe PreE in singletons. Generating this signature through algorithmic feature selection differed from the methods we reported previously (21), and was specifically applied to boost classification performance, enhance model efficiency, mitigate overfitting, and increase robustness relative to a profile encompassing a larger set of differentially modulated transcripts (36, 57).

Several observations from this analysis warrant comment. Firstly, we found that complementing differential transcript abundance analysis with molecular stratification scoring permitted a more comprehensive view of the transcriptomics data, revealing insights that might otherwise be obscured. For example, by conventional groupwise analysis, the No PreE DT (Group 2) placentas appeared similar to the normotensive SG (Group 1) reference (differing by 27 transcripts at FDR<0.05 and with a similarity index of 0.90); however, molecular classification signature scoring revealed that several of the individual samples possessed more PreE-like transcriptional profiles, even in the absence of clinical manifestations. Additionally, the No PreE MT (Group 3) showed an unexpectedly high similarity to the early-onset PreE in singletons (Group 7) by expressing PreE-like gene and pathway patterns. One possible explanation is that the PreE gene expression pathways may become activated prior to the onset of the clinical manifestation. This observation requires further investigation because the limited number of MT specimens did not allow us to generate a full characterization of the relationship between molecular phenotype and clinical PreE in monochorionic gestations.

Furthermore, we observed transcriptional discordance among DT sibling placentas, particularly in those from pregnancies complicated by PreE. Interestingly, disconfirming our expectations, the degree of intertwin transcriptional divergence did not differ between the normotensive and the PreE groups. When the 98-transcript molecular classifier was used to cluster the twin samples, we determined that 57% (8/14) individual DT No PreE (Group 2) samples were within the 80% CI of the No PreE centroid with just one sample falling inside the PreE centroid (Fig.2E). Given that one Group 2 sample (T08A) fell within the PreE centroid CI while many more of the twin No PreE samples clustered between the centroids, it is possible that, at the time of birth, some asymptomatic twin patients were in the process of transitioning toward clinical PreE manifestations. This hypothesis is supported by prior work showing that imbalances in circulating angiogenic factors often preceded the clinical manifestation of maternal disease, including PreE, in twin pregnancies (58). We also found that 90% (9/10) of the DT PreE (Group 4) specimens were clustered between the centroid CIs in the region occupied by a majority of the new singleton (Group 6) samples. Since the classification signature was developed using extremes of clinical phenotypes, we anticipated that samples from the expanded spectrum of PreE manifestations would fall within this intermediate range. We also predicted that relative to singletons with PreE, twins with PreE would show fewer changes consistent with established patterns of pathway dysregulation associated with this condition. Though the isolated functional analyses were consistent with this interpretation (see Figs. 4&5), in the aggregate, roughly half (8/14) of the individual PreE twin placentas, as well as some from the normotensive group (T08 and T16), displayed perturbations recapitulating patterns characteristic of SG PreE samples.

In studies of twin pregnancies with suspected PreE, alterations in circulating angiogenic factors beyond that associated with twinning alone predicted subsequent adverse outcomes (52, 59, 60). Conversely, reduced sFlt-1/PlGF ratios precluded the need for imminent provider-initiated delivery (61). Since elevated sFlt-1 in this setting may reflect placental dysfunction (56), we were particularly interested in examining whether *FLT1* mRNA abundance increased relative to most placental transcripts. A novel finding was that the Singscore ES and Mahalanobis distance metrics based on the molecular classification signature were well correlated with birthweight, placental *FLT1*, *ENG*, and the *FLT1*/*PGF* ratio, which are well established markers of symptomatic and asymptomatic PreE in both singletons and multiple gestations (61, 62). Therefore, the confidence in the validity of our scoring system remained robust. Although recent work in SG pregnancies with PreE showed that circulating sFlt-1/PlGF was significantly correlated with placental sFlt-1 protein abundance (63), we recognize that the relationship between placental *FLT1* mRNA and truncated sFlt-1 protein expression is complex, requiring alternative splicing and/or proteolytic cleavage events (64, 65). Nonetheless, the present results are consistent with the report of Nevo et al., which identified elevated *FLT1* mRNA and sFlt-1 protein expression that was discordant between DT placentas in the setting of PreE (66). These results favor an interpretation whereby the pathomechanisms of PreE in twins transcend the risks stemming from excess placental mass alone and, furthermore, may involve the twin placentas unequally.

Unlike the more abundant transcriptional profiling studies of SG placentas in PreE, those involving twin placentas in this context have been limited. For example, Leseva et al. performed placental microarray profiling in a small sample of DT pregnancies (2 normotensive and 2 with PreE) and identified 5 transcripts (*CYP1B1*, *DEFA1B*, *MIR1973*, *MIR4307HG*, and an uncharacterized transcript on chromosome 19) associated with PreE (67), none of which were identified as differentially abundant in our DT PreE samples (Table S2). However, several studies that have interrogated twin placentas with discordant fetal growth or selective fetal growth restriction (FGR) may provide insights into expression signatures associated with impaired placental function and help to contextualize the present results. For example, in DTs with discordant growth, Roh et al. demonstrated altered placental expression of several hypoxia-regulated transcripts (*CCN2*, *CSH1*, *FSTL3*, *NDRG1*, *PHLDA2*, *PLIN2*, *VEGFA*) in the growth-restricted twins (68). Additionally, Okamoto and colleagues also observed upregulation of *FSTL3* in addition to *IGFBP1* in a growth-restricted DT placenta (69), while Biesiada et al. identified differential placental expression *ANGPT2*, *GLIS3*, *KLF4*, and *LEP* among discordant DT pairs (70). These studies are consistent with the intertwin placental transcriptional discordance we observed.

## Strengths and Limitations

The strengths of our study include the thoroughness of our analytical approach and our attempts to avoid reliance on single metrics for molecular classification. Limitations of the current work include a relatively small sample size, the cross-sectional study design (which limits causal inference), the limited regional sampling of each placenta (since tissue heterogeneity may exist and variability within each placenta may not be adequately sampled), and the inability to assess whether an individual pregnancy was on a trajectory towards developing a hypertensive disorder at the time of delivery.

Furthermore, only one high-dimensional profiling modality was assessed in the present work, and we were unable to relate our observations to associated metabolomic, epigenetic, and proteomic changes in the same tissues. We endeavored to compensate for the small sample size through careful specimen selection from a large biorepository.

## Perspectives and Conclusion

In conclusion, the current study aimed to investigate molecular differences between placentas of twin pregnancies at the transcriptional level. It provides new evidence relevant to the development of nucleic acid-based prenatal testing techniques, such as cell-free DNA and RNA, to detect the risk of developing PreE in its asymptomatic phase. These should be accompanied by the pursuit of preventative therapies capable of halting the disease’s progression.

## Author Contributions

Designed research: WEA, CSB, and IAB conceived and developed the study design. Performed research: CSB and IAB participated in patient recruitment, abstraction of clinical data, and the collection and processing of biological specimens; GZ executed the RNA extraction from specimens. Analyzed data: WEA and IAB conducted the data analysis with input from CSB and TLB. Wrote the paper: WEA wrote the first draft of the manuscript with input from TLB, CSB, and IAB. All authors participated in reviewing the work and contributed with critical review and revisions to the drafts of the manuscript.

## Data Availability

https://www.ncbi.nlm.nih.gov/geo/query/acc.cgi?acc=GSE272342

## Acknowledgments

Portions of the analysis presented were conducted using the University of Illinois Chicago Advanced Cyberinfrastructure for Education and Research high-performance computing resources. The authors gratefully acknowledge the Nucleic Acid Shared Resource at The Ohio State University Comprehensive Cancer Center for technical support while this work was ongoing. Portions of this work were presented in abstract form at the 39^th^ (February 11 - 16, 2019, Los Vegas, NV), 40^th^ (February 3 - 8, 2020, Grapevine, TX), and the 44^th^ (February 10 -14, 2024, National Harbor, MD) Annual Pregnancy Meetings of the Society for Maternal-Fetal Medicine.

## Supplement

**Tables S1–5.**

**Figures S1-8.**

**Supplemental Tables**

**Table S1. Detailed clinical characteristics for the present study.**

**Table S2. Pairwise comparisons for full DESeq2 model.**

**Table S3. Pairwise comparisons for subset DESeq2 models.**

**Table S4. Transcripts included in preeclampsia molecular classification signature.**

**Table S5. Summary and data for transcript-based placental scoring metric regression models.**

---

**Figure S1.**
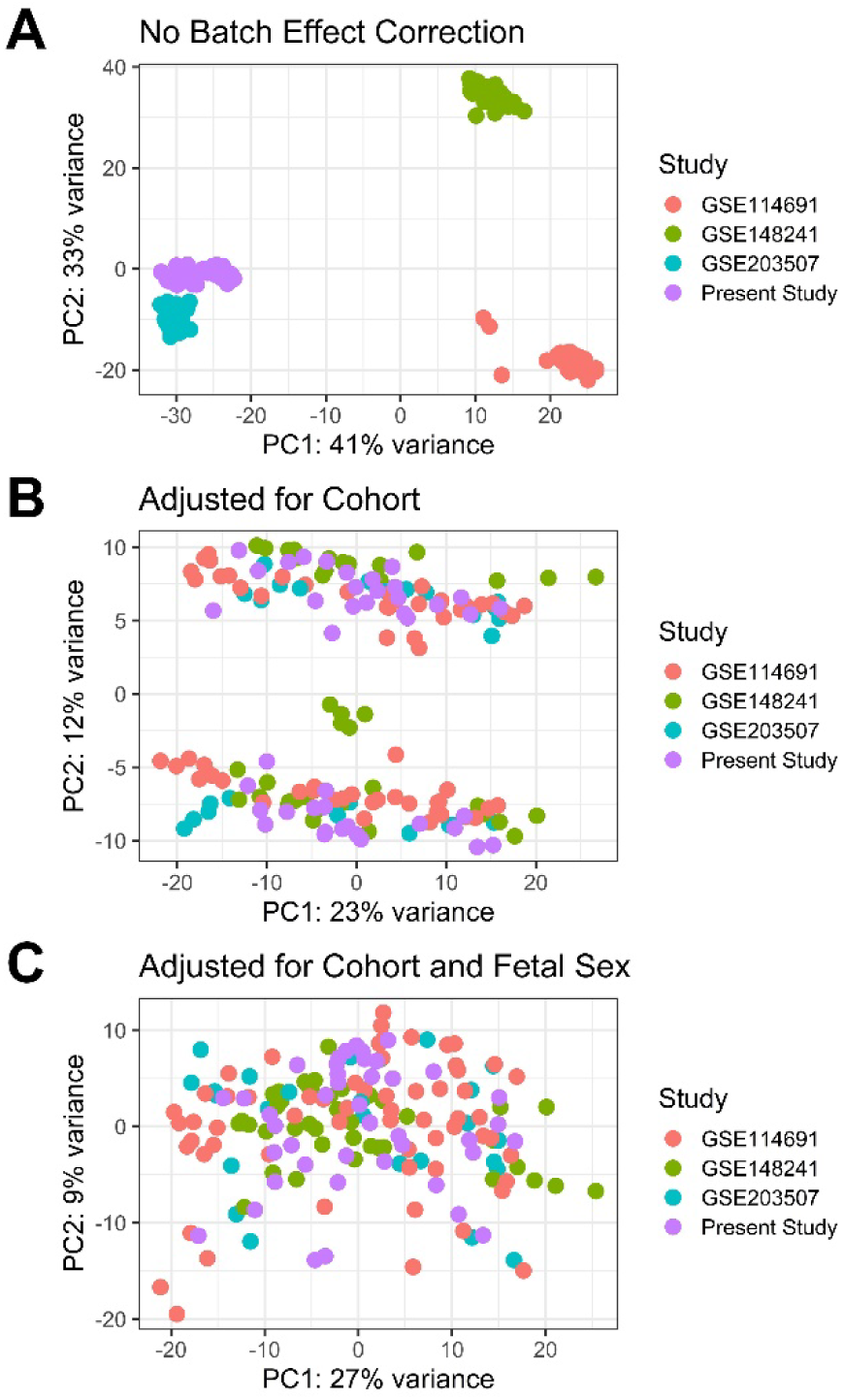
Effects of batch correction on samples from all four RNA-seq cohorts. (A) Principal component (PC) analysis for the RNA-seq samples prior to batch correction. (B) PC analysis for the RNA-seq samples following correction for study using the removeBatchEffect function in the limma R library. (C) PC analysis for the RNA-seq samples following correction for study and fetal sex using the removeBatchEffect function in the limma R library. Data for plots were calculated with the plotPCA function in the DESeq2 R package using the top 500 transcripts selected by highest row variance.

**Figure S2.**
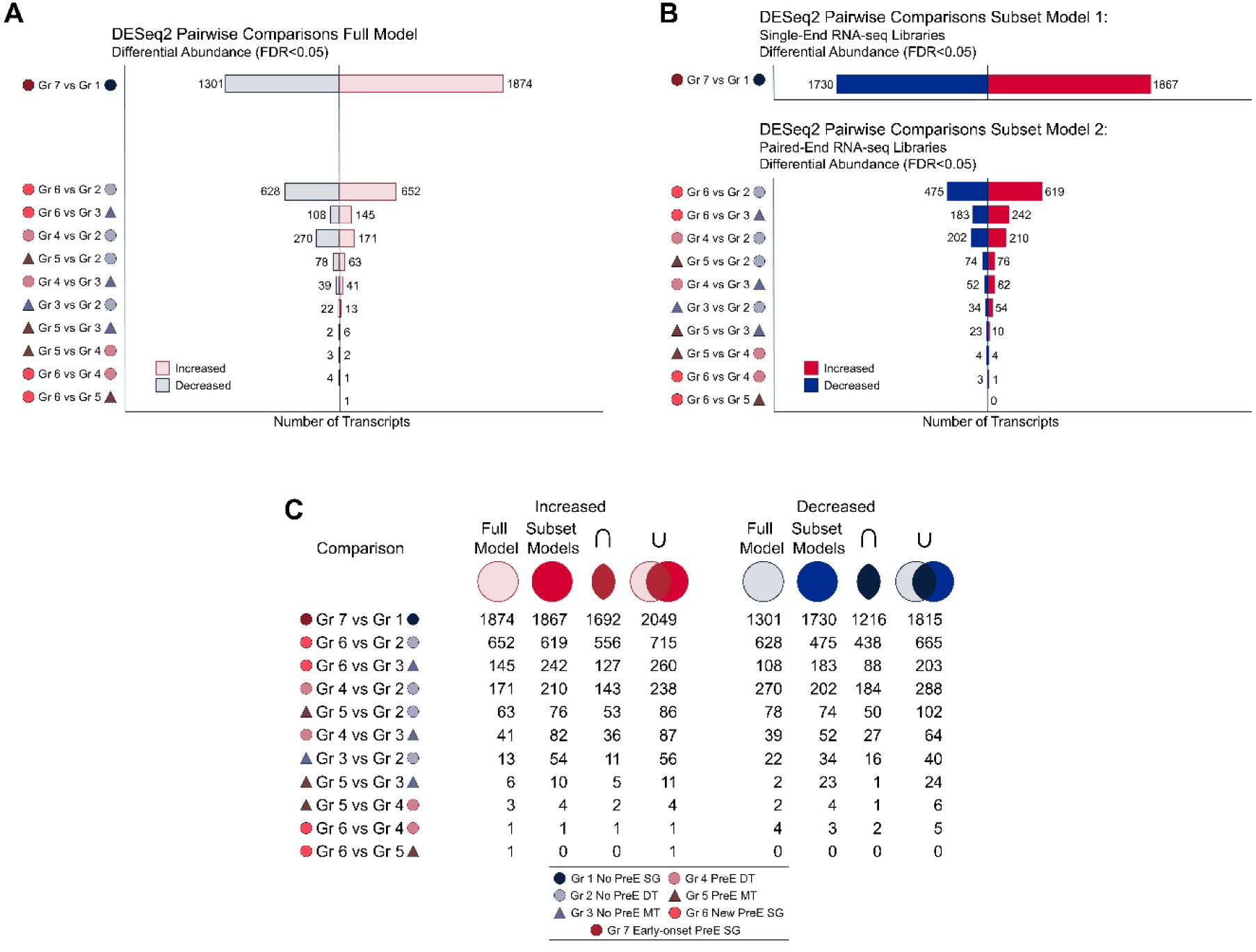
Sensitivity analysis for differential abundance models. For the twins cohort inclusive of the 32 individual twin samples, 10 newly sequenced singleton placentas, and 25 reference samples from GSE203507 (15 PreE specimens, 10 controls), sensitivity analysis was performed using RNA-seq statistical models comprising two subsets of the full DESeq2 model, each restricted to samples from the individual batches that differed by library preparation protocol (single-end and paired-end) but were sequenced at a single facility. (A) Stacked bar plot showing pairwise comparisons from the full model that correspond to those of the two subset models (Wald test, FDR<0.05). (B) Bar plots showing pairwise comparisons for the two subset models (Wald test, FDR<0.05). Subset Model 1 comprised the 25 specimens from GSE203507 that were constructed using single-end library reagents, while Subset Model 2 consisted of the 42 remaining samples prepared using pair-end reagents and protocols. (C) Table showing the number of differentially abundant transcripts (Wald test, FDR<0.05) for each comparison in the full and subset models with the intersection and union of the resulting transcript lists. Abbreviations: DT, dichorionic twin; Gr, group; MT, monochorionic twin; PreE, preeclampsia; SG, singleton gestation.

**Figure S3.**
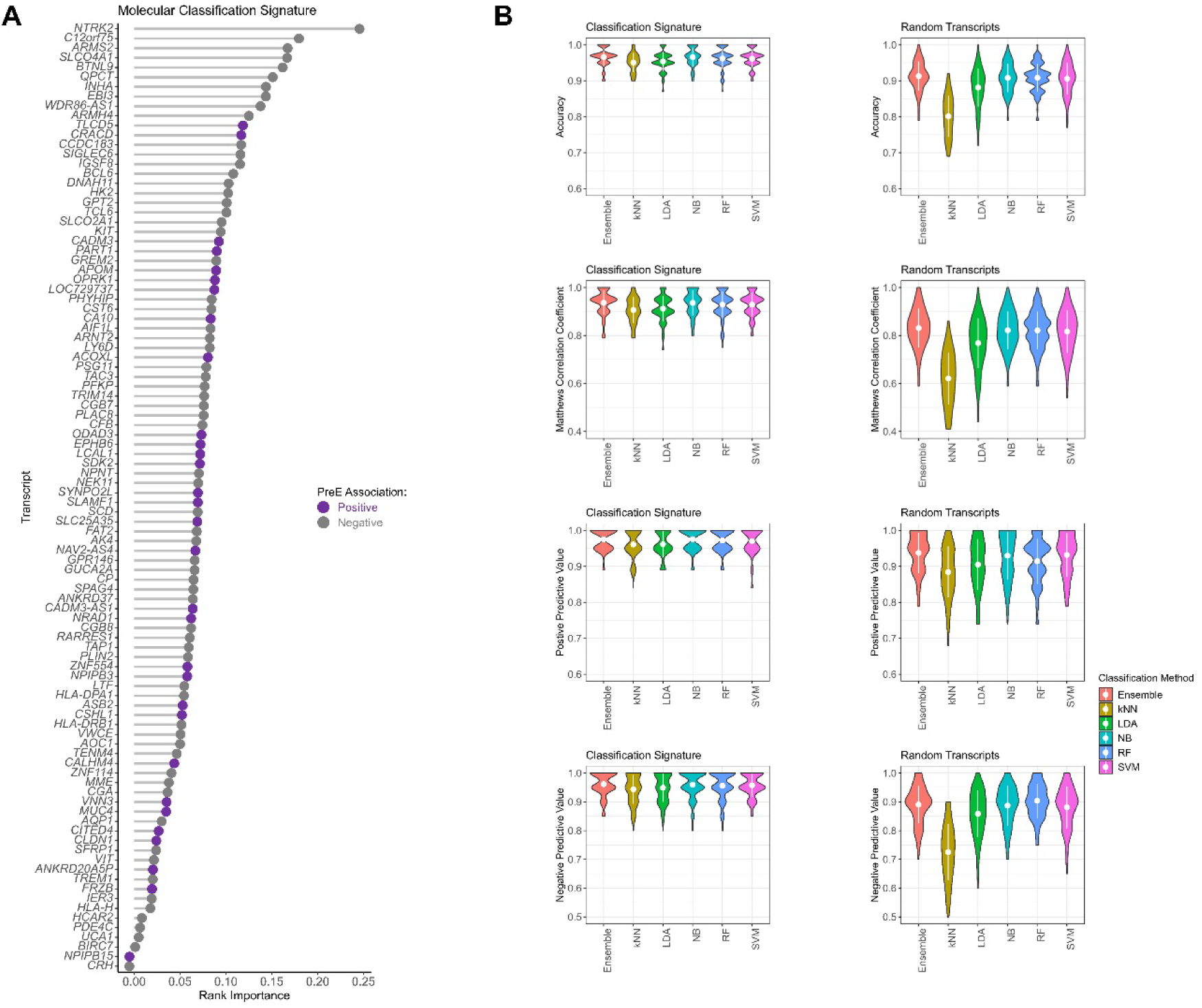
Binary classification accuracy for 98-transcript molecular signature in 3-cohort benchmark PreE transcriptome dataset. The DaMiR.EnsembleLearning function was used to evaluate the robustness of the 98-transcript molecular signature compared to a randomly selected transcript signature of equal size. Bootstrap sampling (70/30 data split) of either dataset was used to generate training and test samples from the original 3-cohort dataset. The training set was then split to train and cross-validate a series of 5 statistical learning classifiers. From these, an Ensemble classifier was generated using a weighted majority voting approach involving a linear combination of the weights and predictions of the individual classifiers. The final performance was then assessed using the original samples held over for testing. The process was repeated over 100 iterations. (A) Dot plot (adapted from the ggdotchart function in the ggpubr R library and rendered using ggplot2) ranked by feature importance for 98-transcript classification signature. Transcripts positively associated with PreE are represented in purple, while those negatively associated with PreE are shown in gray. Feature importance was calculated using regressional ReliefF. (B) Violin plots showing the performance of the classification signature (left panels) relative to that of a panel of 98 randomly selected transcripts (right panels) for samples in the 3-cohort dataset across 5 supervised binary classification algorithms. Averaged performance metrics and standard deviations are represented by white dots and lines, respectively. Abbreviations: kNN, k-nearest neighbor; LDA, linear discriminant analysis; NB, naive Bayes; PreE, preeclampsia; RF, random forest; SVM, support vector machine.

**Figure S4.**
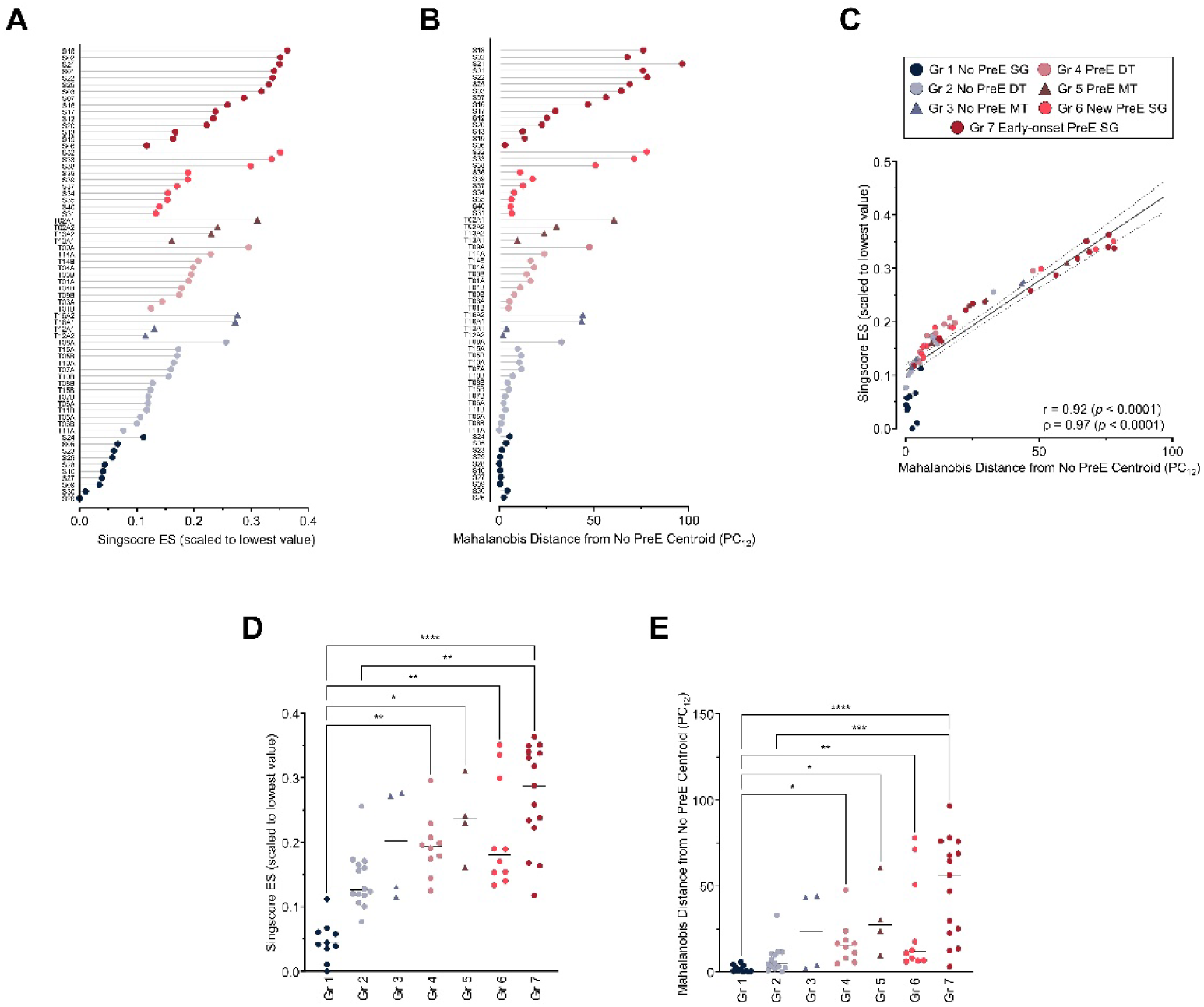
Application of the PreE molecular classifier for quantification of the placental PreE spectrum. Distances along the placental PreE spectrum were calculated based on the 98-transcript PreE molecular classification signature. (A) Dot plot Singscore ES (scaled to the lowest value calculated among the samples) for singleton and twin samples (see also Figure S7 for individual twin pairs). (B) Dot plot Mahalanobis distance for SG and twin samples. Mahalanobis distance was calculated from the control (No PreE) centroid shown in Fig. 1 along the first two principal components (PC_12_) following PCA using the 98-transcript PreE molecular classification signature. Dot plots were adapted from representations made using the ggdotchart function from the ggpubr R package and rendered using GraphPad Prism. (C) Scatterplot of Singscore ES versus Mahalanobis distance, shown with the correlation coefficients for this relationship (r=0.92, *p*<0.0001; ρ=0.97, *p*<0.0001). Higher scores for the Singscore ES and Mahalanobis distance metrics indicate transcriptomic signatures more consistent with PreE. (D) Plot of scaled Singscore ES (panel A data) showing individual values for each group, with bars representing medians. High-scoring samples have expression signatures that were more consistent with PreE. Group medians were evaluated using the Kruskal-Wallis test (H=39.83, *p*<0.0001). (E) Plot of Mahalanobis distance (data as in B) showing individual values for each group, with bars representing medians. Medians were evaluated using the Kruskal-Wallis test (H=38.2, *p*<0.0001). Asterisks indicate levels of significance in multiple comparisons using the Dunn test: *, *p*<0.05; **, *p<*0.01; ***, *p<*0.001*; ****, p*<0.0001. Abbreviations: DT, dichorionic twin; Gr, group; MT, monochorionic twin; PC, principal component; PreE, preeclampsia; SG, singleton gestation.

**Figure S5.**
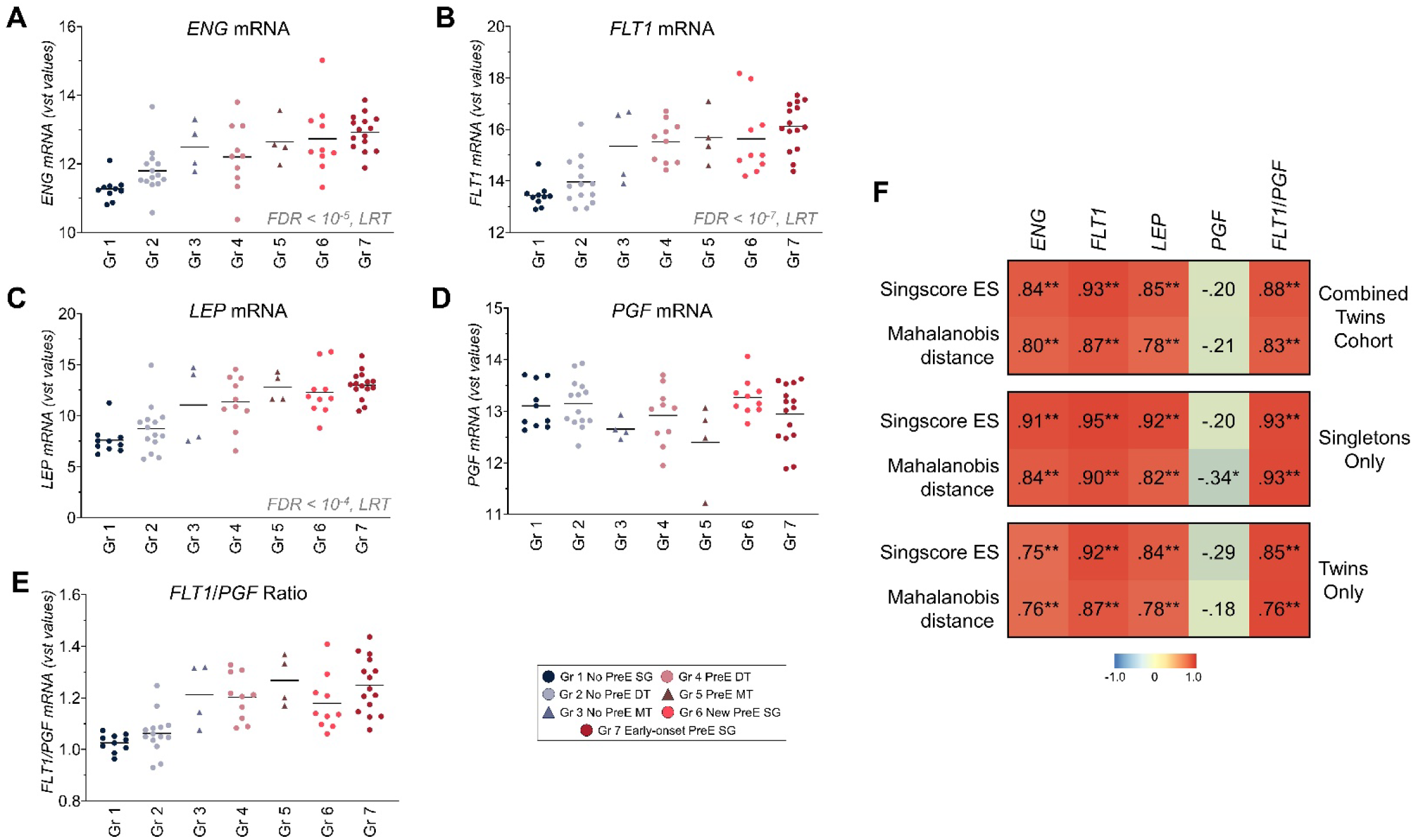
Placental expression of *ENG*, *FLT1*, *LEP*, *PGF*, and *FLT1*/*PGF* transcripts in the twins cohort dataset and their correlations with molecular stratification scores. (A-E) Dot plots showing the batch-corrected, variance-stabilizing transformed, normalized expressions of *ENG*, *FLT*1, *LEP*, and *PGF* transcripts and the ratio *FLT*/*PGF* in samples from individual villous placentas of the twins cohort grouped by clinical features. The expression of *ENG*, *FLT*1, and *LEP* differed significantly (FDR<0.05) among the groups as determined by DESeq2 using the likelihood-ratio test (LRT). (F) Heat maps showing the Pearson correlations between the above transcripts and the Singscore ES and Mahalanobis distance scores for the twins cohort overall (top), the singleton samples only (middle), and the twin samples only (bottom). Singscore ES (scaled to lowest value) and Mahalanobis distance (from the No PreE centroid) metrics were calculated based on the 98-transcript PreE molecular classification signature that did not include *FLT1*, *ENG*, *LEP*, or *PGF*. Asterisks indicate statistical significance levels: *, *p*<0.05; **, *p<*0.0001. Abbreviations: DT, dichorionic twin; ENG, endoglin; FLT1, FMS-related tyrosine kinase 1; Gr, group; LEP, leptin; LRT, likelihood-ratio test; MT, monochorionic twin; PC, principal component; PreE, preeclampsia; PGF, placental growth factor; SG, singleton gestation.

**Figure S6.**
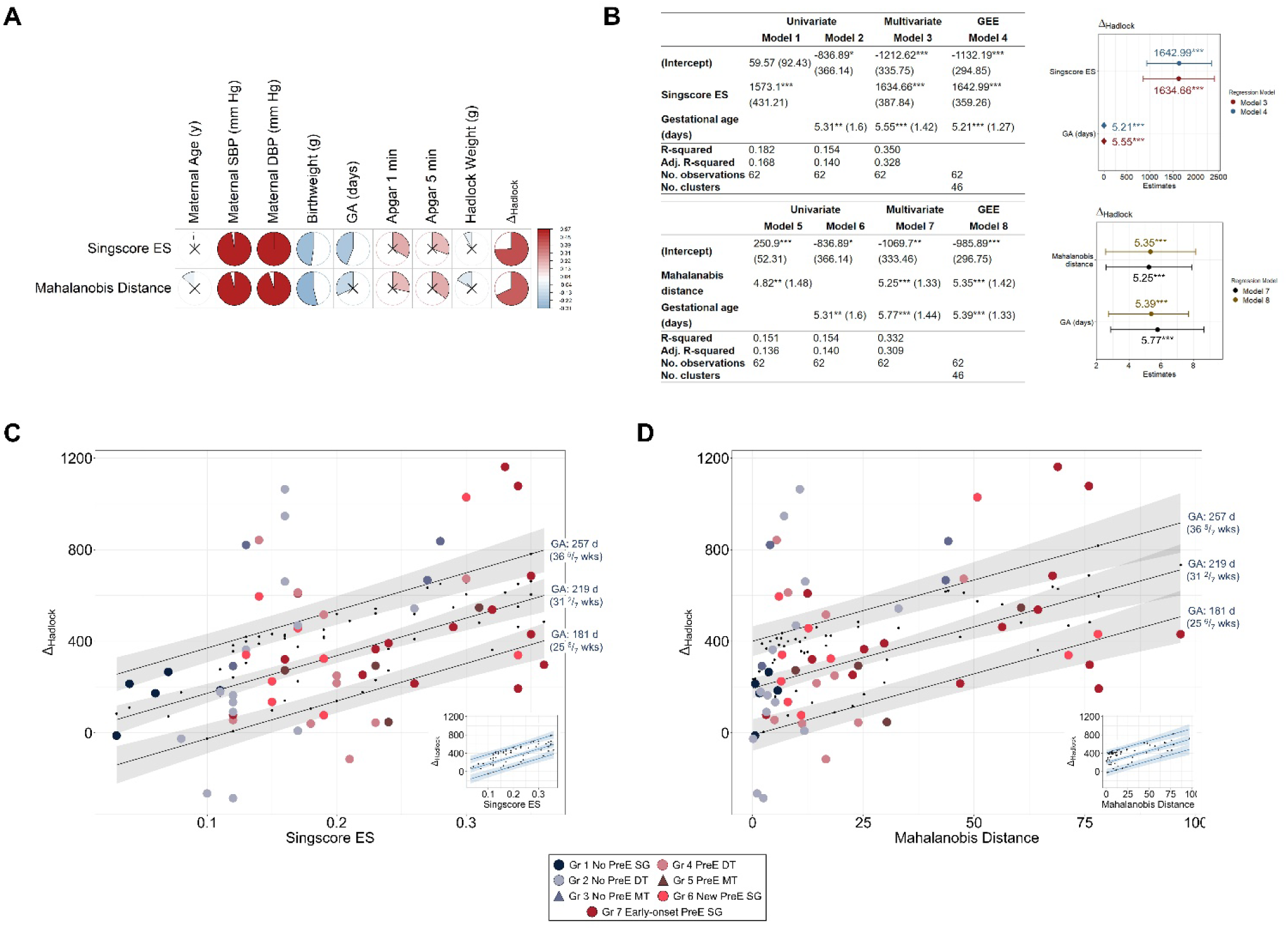
Transcript-based placental scoring metrics are independent predictors of deviation from the expected fetal weight in singleton and twin pregnancies. (A) Pearson correlation matrix (generated with corrplot v0.92) showing the associations between the Singscore ES or Mahalanobis distance scores and selected clinicodemographic variables. The circle areas and shading indicate the size and direction of the correlation coefficients (scaled to minimum and maximum values), while the “X” glyph denotes correlations that were not statistically significant. The statistical significance of the indicated associations was unaltered in mixed effects models in which the pregnancy identifier was included as a random effect to account for non-independence of pregnancy-related characteristics (maternal age, maternal blood pressure, and gestational age at delivery) for the individual twin scoring metrics. (B) Summary of regression models. Simple and multiple linear regression models were used to test whether the Singscore ES and Mahalanobis distance scores significantly predicted deviation from the Hadlock expected fetal weight (Delta, Δ_Hadlock_). Generalized estimating equation (GEE) models (Models 4 and 8 for the Singscore ES and Mahalanobis distance scores, respectively) were also used to account for the paired structure of the twins data; these were performed using the glmgee function in the glmtoolbox v0.1.11 R package with Gaussian variance and an unstructured correlation. The tables summarize the model results, showing the model coefficient estimates, standard errors in parentheses, and asterisks (*,**,***) indicating significance at the 90%, 95%, and 99% levels, respectively. The plots show the coefficient estimates and 95% confidence intervals for the predictor variables in the multiple regression and GEE models, which were similar. Additional details are presented in Table S5. (C) Scatterplot showing predictions from the GEE and multiple regression (inset) models using Singscore ES and gestational age in days as predictor variables. The larger, colored circles represent the raw data, while the fitted values are depicted by small black circles. Fitted regression lines with standard errors for the predicted Δ_Hadlock_ values are shown for three fixed gestational age values across a range of Singscore ES values. (D) Scatterplot of GEE model predictions as in panel C, but for the Mahalanobis distance scores. The plots in panels C and D were adapted from the ggPredict function of the ggiraphExtra v0.3.0 package and display the GEE model estimates, the results of which were similar to those of the multiple regression model estimates (Models 3 and 7, insets). Abbreviations: d, days; DT, dichorionic twin; DBP, highest maternal diastolic blood pressure; GA, gestational age; Gr, group; MT, monochorionic twin; PreE, preeclampsia; SG, singleton gestation; SBP, highest maternal systolic blood pressure; wks, weeks.

**Figure S7.**
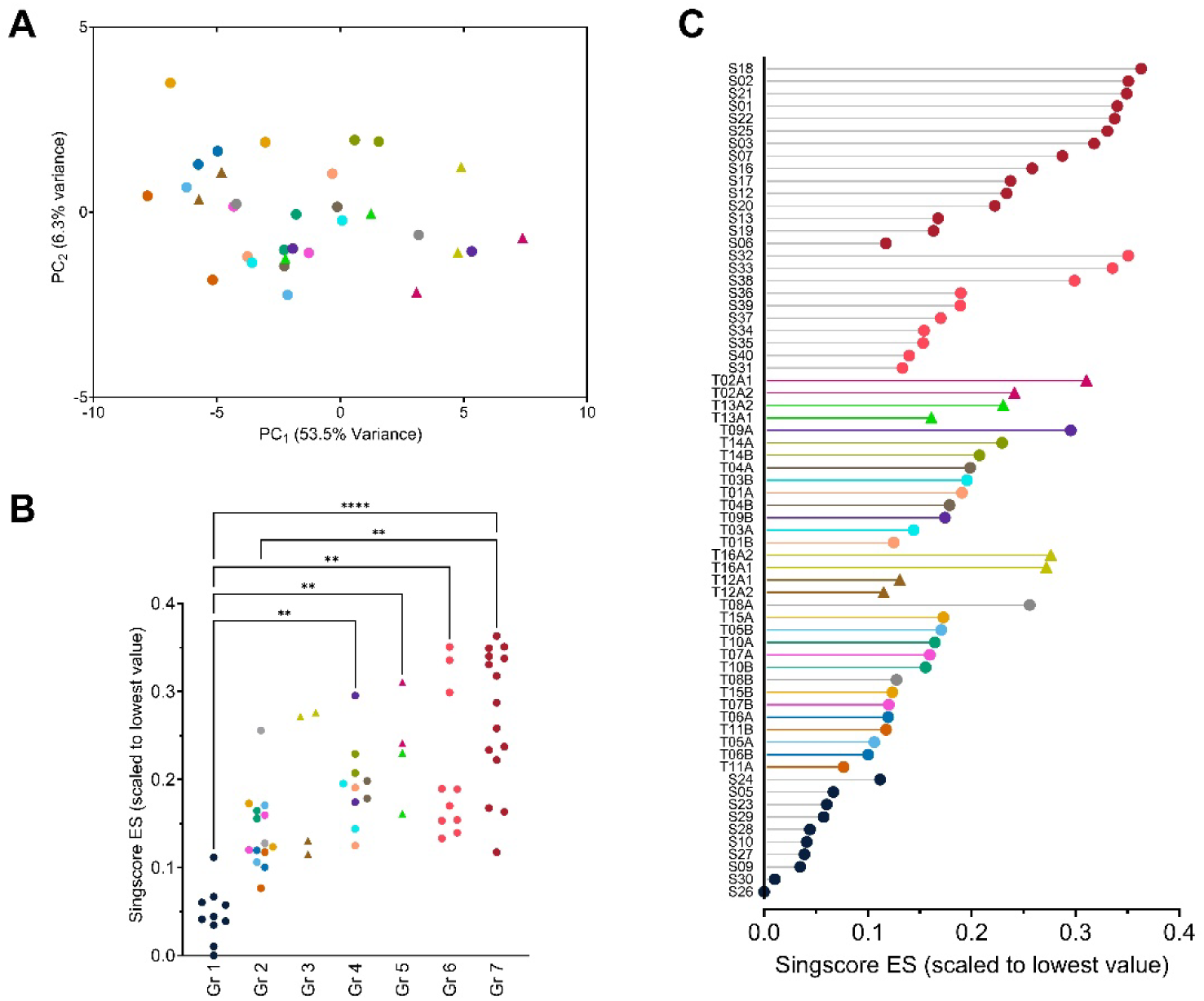
Recolored depictions of selected figures to highlight positions of twin pairs. Each of the individual twin pairs is represented with a unique color. (A) Distribution of twin placenta samples within PC_12_ coordinates as in Figure 2E. (B) Plot of Singscore ES (scaled to the lowest calculated value among the samples) showing individual values for each group as in Figure 2F. (C) Dot plot Singscore ES (scaled to the lowest value calculated among the samples) for singleton and twin samples as in Figure S4A. Abbreviations: ES, expression score; Gr, group.

**Figure S8.**
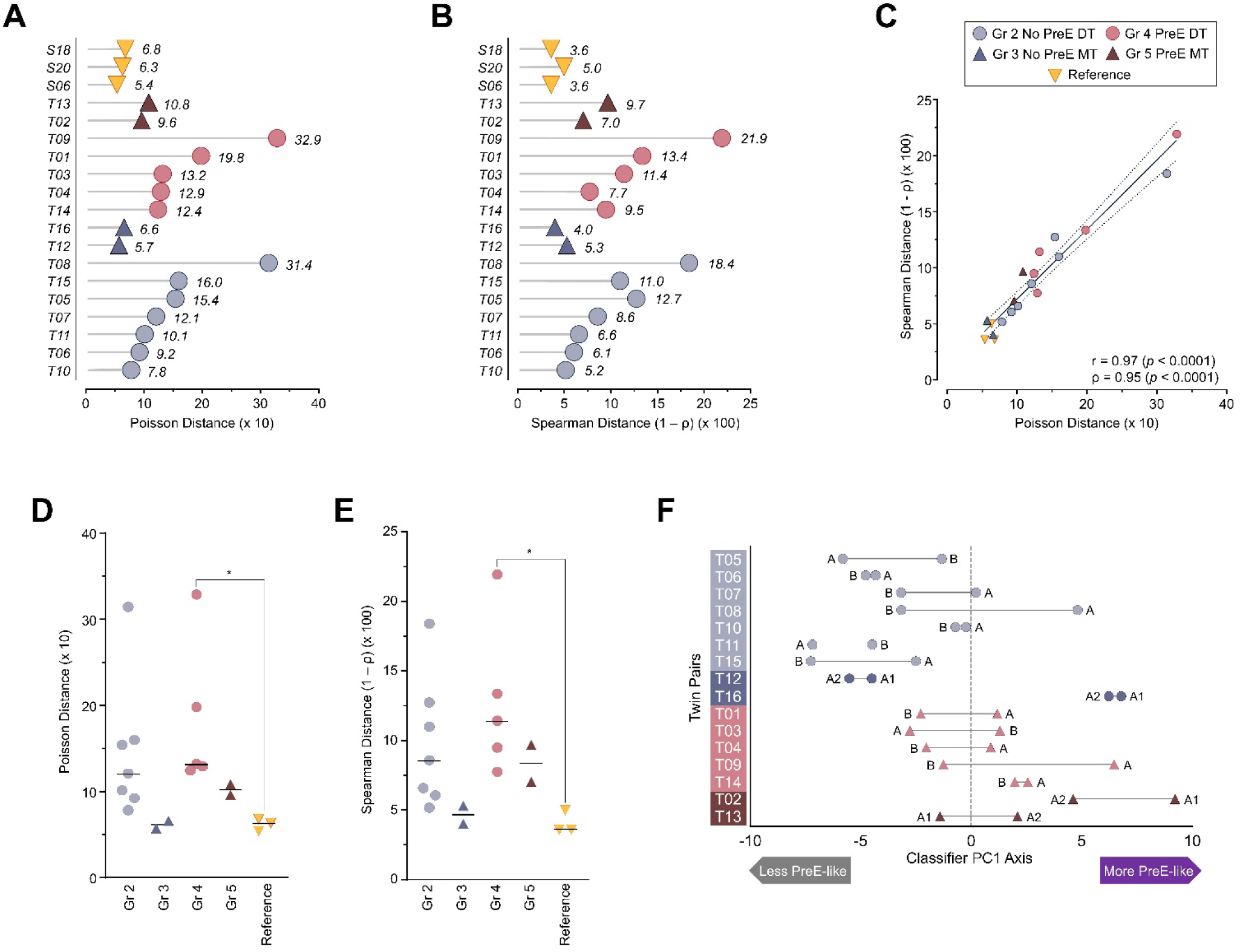
Distance between paired sibling twin placentas. Using the 98-transcript PreE molecular classification signature, we calculated distances between twin sibling placental sample pairs using Poisson dissimilarity and Spearman correlation distance (1 - ρ). As a reference, we calculated the distances between the singleton placental samples (all from cases with severe early-onset PreE) that were extracted and sequenced both in the present study and the prior GSE203507 dataset. (A) Dot plot of Poisson distance for the twin samples and the singleton reference specimens. (B) Dot plot of Spearman correlation distance for the samples in panel A. Distances were scaled to provide values within a consistent range to aid comparison. For both metrics, increasing distance indicates a greater degree of between-sample dissimilarity. Dot plots were adapted from representations made using the ggdotchart function from the ggpubr R package and rendered using GraphPad Prism. (C) Scatterplot of Poisson distance versus Spearman correlation distance shown with the correlation coefficients for this relationship (r=0.97, *p*<0.0001; ρ=0.95, *p*<0.0001). (D) Plot of Poisson distance (data as in panel A) showing individual values for each group, with bars representing medians. Medians were evaluated using the Kruskal-Wallis test (H statistic 12.4, p=0.015). (E) Plot of Spearman correlation distance (panel B data) showing individual values for each group, with bars representing medians. Medians were evaluated using the Kruskal-Wallis test (H statistic 11.3, *p*=0.024). Asterisks indicate p<0.05 (Dunn test). (F) Linked dot plots showing relative positions of twin pair values along the first principal component axis following application of the 98 transcript molecular classifier. Abbreviations: DT, dichorionic twin; ES, expression score; Gr, group; MT, monochorionic twin; PreE, preeclampsia.

**Figure S9.**
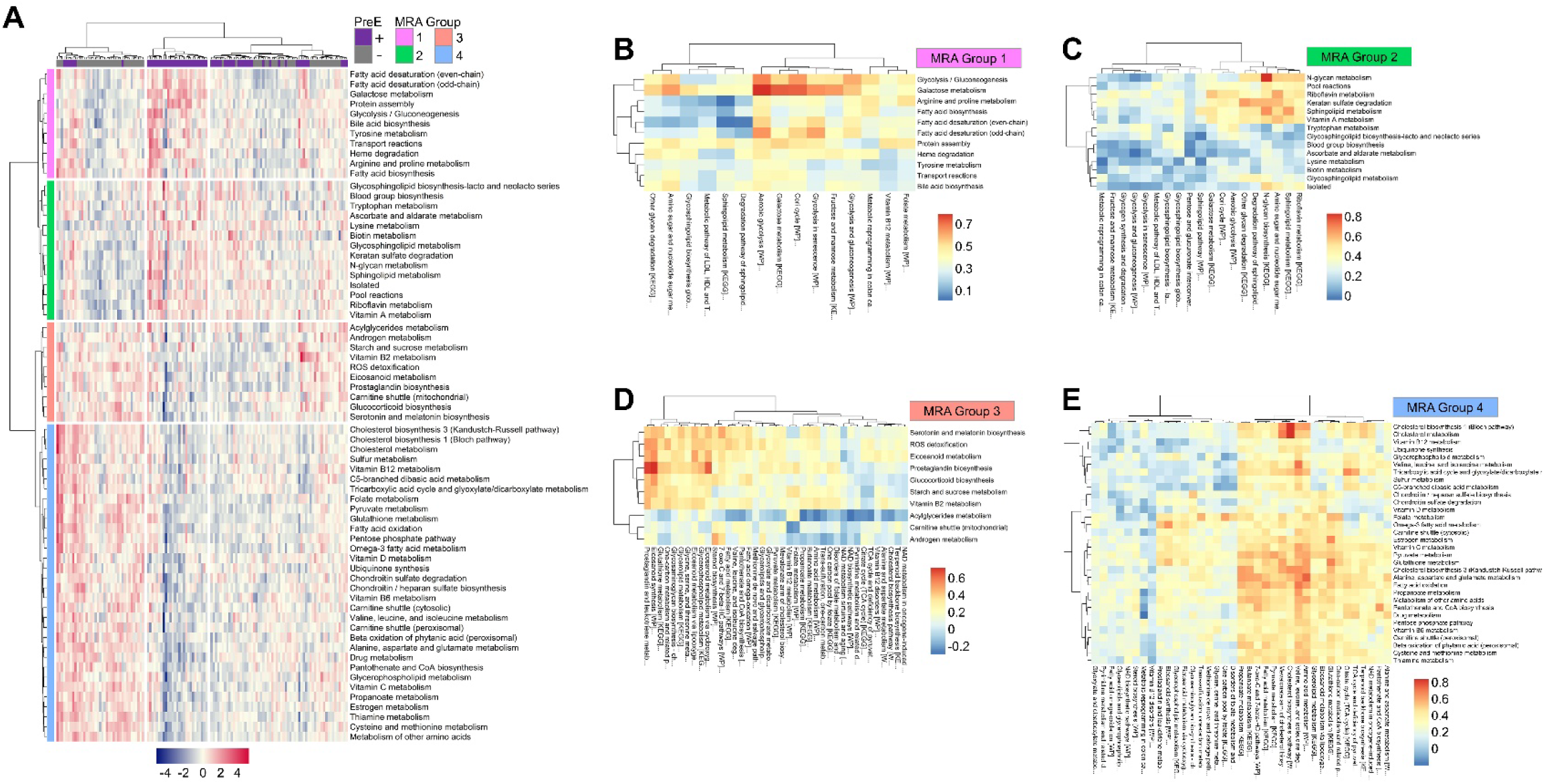
Metabolic reaction activity (MRA) scoring for individual samples in the 3-cohort benchmark PreE dataset. Pathway-level MRA scores were calculated with METAFlux software for 13,082 cataloged reactions using the batch-corrected and variance-stabilizing transformed expression data (25,289 transcripts) of the 3-cohort PreE dataset (n=127 samples). (A) Clustered heat map of pathway-level metabolic MRA scores. The PreE villous placenta specimens were differentiated from the No PreE singletons by 67 MRA scores (FDR<0.05, Mann–Whitney test with *p*-value adjustment). The MRA scores (rows) were segregated into 4 major groups by hierarchical clustering, with the individual samples subdividing into 3 principal column clusters. (B-E) Heat maps showing Spearman correlations between MRA scores and corresponding GSVA scores (metabolism-related KEGG and WIkiPathway gene sets) for individual samples in the 3-cohort dataset for each of the 4 MRA score groups shown in panel A. Abbreviations: DT, dichorionic twin; Gr, group; GSVA, gene variation analysis; KEGG, Kyoto Encyclopedia of Genes and Genomes; MT, monochorionic twin; MRA, metabolic reaction activity; PreE, preeclampsia; SG, singleton gestation; WP, WikiPathway.

